# *In Vivo* Topographic Associations Between Tau Pathology, Atrophy, and Symptom Domains in Patients with Progressive Supranuclear Palsy

**DOI:** 10.1101/2025.06.23.25329885

**Authors:** Yuko Kataoka, Ryoji Goto, Yoshikazu Chishiki, Kenta Osawa, Asaka Oyama, Hideki Matsumoto, Masanori Ichihashi, Sho Moriguchi, Yuki Momota, Tetsuji Kamada, Chie Seki, Kiwamu Matsuoka, Kosei Hirata, Shin Kurose, Masaki Oya, Naomi Kokubo, Hitoshi Shinotoh, Hitoshi Shimada, Kazunori Kawamura, Ming-Rong Zhang, Takahiko Tokuda, Keisuke Takahata, Kenji Tagai, Makoto Higuchi, Hironobu Endo

**Affiliations:** Advanced Neuroimaging Center, Institute for Quantum Medical Science, National Institutes for Quantum Science and Technology (QST), Chiba, Japan; Department of Advanced Nuclear Medicine Sciences, Institute for Quantum Medical Science, QST, Chiba, Japan; Department of Oral and Maxillofacial Radiology, Tokyo Dental College, Tokyo, Japan; Department of Functional Neurology & Neurosurgery, Center for Integrated Human Brain Science, Brain Research Institute, Niigata University, Niigata, Japan; Department of Neuroetiology and Diagnostic Science, Osaka Metropolitan University Graduate School of Medicine, Osaka, Japan

## Abstract

**Background and Objectives:** Progressive supranuclear palsy (PSP) is a four-repeat tauopathy characterized by subcortical atrophy and distinctive motor impairments. Although tau pathology is thought to drive neuronal loss, the spatial relationships among tau deposition, regional atrophy, and domain-specific symptoms remain incompletely understood. We aimed to determine how the topographic distribution of tau pathology and regional brain atrophy relate to distinct motor symptom domains in PSP and to clarify their contributions to clinical manifestations.

**Methods:** A single-center, cross-sectional study was conducted. Patients with PSP were recruited after diagnosis at neurology departments and were referred to our institution. All PSP cases underwent neurological assessments, including PSP Rating Scale. Tau positron emission tomography (PET) imaging with florzolotau (18F) and T1-weighted magnetic resonance imaging were obtained from patients and healthy controls to assess associations between domain scores, tau accumulation, and regional brain volumes. Path analysis was used to examine whether cross-sectional associations were consistent with hypothesized relationships among tau deposits, regional atrophy, and symptom domains.

**Results:** Fifty-eight patients with PSP (mean age, 71.6 years; 43.1% female; Richardson’s syndrome, n = 43) and 52 healthy controls (mean age, 60.8 years; 45.3% female) were included. Subcortical tau accumulation and atrophy demonstrated significant spatial overlap and were associated with motor deficits. “Ocular motor” domain scores were associated with tau deposition in the midbrain tegmentum and globus pallidus. “Gait and midline” domain scores were associated with tau in the globus pallidus, thalamus, and subthalamic nucleus. “Limb motor” domain scores were associated with tau in the primary motor and somatosensory cortices, as well as atrophy in the angular, supramarginal, and temporal cortices. These findings were identified at a family-wise error corrected cluster-level threshold of *p* < 0.05. Path analysis indicated that tau in subcortical structures was associated with local neuronal loss and with impairments in the “ocular motor” and “gait and midline” domains.

**Discussion:** High-contrast tau imaging clarifies the neuropathological basis of key symptoms and underscores distinct tau-induced neurotoxic effects in subcortical versus neocortical regions. Specifically, tau deposition in the neocortex may contribute to “limb motor” impairments through local effects and potentially through more distributed neurodegenerative processes. These findings support anatomically distinct tau–neurodegeneration pathways underlying key PSP symptom domains.

## Introduction

Progressive supranuclear palsy (PSP) is an idiopathic neurodegenerative disorder lacking effective treatments. Clinically, it manifests with motor impairments resembling Parkinson’s disease, alongside distinctive features like ocular motor dysfunction and postural instability. Neuropathologically, PSP is characterized by neuronal and glial inclusions composed of four-repeat tau isoforms, including globose tangles, coiled bodies, and tufted astrocytes. These tau aggregates predominantly affect subcortical structures such as the thalamus, subthalamic nucleus, and brainstem, although neocortical regions, particularly in the frontal lobe, are also frequently involved.^1–4^ Accordingly, PSP is classified within frontotemporal lobar degeneration with tau pathology (FTLD-Tau).^5^

A hallmark of PSP in morphometric neuroimaging is midbrain atrophy, commonly recognized as the hummingbird sign,^6^ correlating with neurological deficits assessed by the PSP Rating Scale.^7^ Tau deposits are thought to contribute to neuronal loss, particularly in the brainstem. However, the extent to which tau accumulation leads to regional volume loss through direct or network-mediated effects remains unclear. Investigating these processes in vivo could provide crucial insights into the pathological basis of symptomatic manifestations. While animal models expressing four-repeat tau isoforms exhibit neuronal loss,^8^ their ability to replicate PSP-specific neurotoxic mechanisms remains uncertain. Moreover, while PSP-derived tau fibril inoculation in rodents induces neuronal and glial tau pathologies,^9^ prominent neuronal loss leading to measurable atrophy has not been observed.

Given these discrepancies, it is crucial to establish links among tau aggregation, neuronal loss, and functional decline using human data. Although postmortem studies have delineated tau inclusions in PSP brains,^4,10^ they do not capture antemortem symptoms, underscoring the value of in vivo assessments to explore spatial relationships between tau pathology, brain atrophy, and symptom severity.

Recently, we developed a tau positron emission tomography (PET) radioligand, florzolotau (18F), which exhibits high contrast for various tau pathologies, including those in PSP.^11^ Florzolotau (18F) enabled in vivo visualization of tau deposits in the thalamus, subthalamic nucleus, basal ganglia, midbrain, and motor cortices in PSP patients,^11,12^ findings align with neuropathological observations. Two independent studies have demonstrated a strong correlation between florzolotau (18F) retention in the subthalamic nucleus and/or midbrain and total PSP Rating Scale scores.^11,13^

Extending these findings, a recent study applied a subtype/stage inference algorithm to florzolotau (18F) PET data and identified distinct progression patterns of tau deposition in PSP, underscoring the potential of machine learning to capture disease heterogeneity and temporal dynamics.^14^ Despite these results, large-scale PET investigations are warranted to elucidate the evolution of tau pathology and its impact on key clinical domains, such as impaired eye movements and gait abnormalities.

Previous studies using flortaucipir (18F) examined tau pathology and brain atrophy in PSP patients across various symptomatic subtypes,^15^ but did not directly examine associations with motor deficits. Furthermore, an imaging-pathology correlation analysis suggested flortaucipir (18F) has limited sensitivity for four-repeat tau aggregates in FTLD,^16^ questioning its utility for staging PSP.^17^ Several studies have evaluated the visual assessment of 4 repeat tauopathies using this tracer,^18,19^ and others have demonstrated that [¹ F]PI-2620 retention in the globus pallidus correlates with disease severity in PSP.^20^ Compared with [¹ F]PI-2620, florzolotau (18F) enables visualization of tau deposition across a broader range of brain areas,^21^ including the midbrain, pons, subthalamic nucleus, thalamus, and precentral cortex, consistent with the distribution of tau inclusions confirmed by postmortem assays. Notably, our recent neuropathological investigation of a PSP case who had undergone PET scans has further supported the utility of this radiotracer for quantitative assessment of 4-repeat tau inclusions across diverse anatomical structures.^22^

Leveraging the ability of florzolotau (18F) to capture tau aggregates in region-specific patterns, the present study aimed to identify the spatial distributions of tau pathology and regional brain volume changes associated with key symptomatic domains in a PSP cohort. By examining concordance and discordance between florzolotau (18F) PET and volumetric MRI, we seek to determine whether four-repeat tau accumulation induces neurotoxicity locally or via network-mediated pathways. Furthermore, path analysis, previously used to test consistency with hypothesized models,^23,24^ was applied to cross-sectional data to estimate associations between disease duration strata, tau-induced neuronal damage associated with symptom categories.

## Methods

### Study Design

This single-center, cross-sectional observational study was conducted at the National Institutes for Quantum Science and Technology (QST), Japan, between January 2018 and May 2023.

### Participants

Patients with PSP were referred to QST after undergoing comprehensive neurological evaluations and receiving a clinical diagnosis at medical institutions with neurology departments.

The diagnosis of PSP was based on the International Parkinson and Movement Disorder Society clinical criteria.^25^ Only patients meeting the criteria for *PSP* or a more definitive diagnostic category were included.

Healthy controls (HCs) were recruited from an institutional volunteer registry maintained at QST. Eligibility criteria for recruitment included the absence of a history of head injury requiring hospitalization, no prior diagnosis of organic intracranial disease, and no ongoing treatment for serious medical conditions. HCs were defined as individuals aged ≥35 years with no history of neurological impairment, a Mini-Mental State Examination score of ≥28, and either a Geriatric Depression Scale score of ≤5 or no history of depression.

Disease severity was assessed using the PSP Rating Scale,^26^ which comprises the following domains: history, mentation, bulbar function, ocular motor function, gait and midline stability, and limb motor function. The PSP Rating Scale was administered and scored by board-certified neurologists with extensive clinical experience. Frontal lobe function was additionally evaluated using the Frontal Assessment Battery (FAB).^27^ For FAB analyses, patients with PSP who had missing FAB scores (n = 3) were excluded, and analyses were conducted using the available data.

All participants were recruited from a dedicated volunteer registry. Aβ-positive cases were excluded based on visual assessment of [^11^C]Pittsburgh Compound-B ([^11^C]PiB) PET scans, independently reviewed by at least three experienced PET specialists.^28^ Details regarding case exclusions are provided in the supplementary figure (eSAP 1) and the trial participant flow diagram.

### MRI Studies

Structural MRI was performed using 3 T MAGNETOM Verio or Prisma scanners (Siemens Healthcare) with a T1 weighted sequence. These images were used for PET coregistration and tissue segmentation in region based volumetric analyses. Detailed acquisition parameters are provided in the eMethods: *MRI Acquisition* section.

### PET Studies

Amyloid pathology was evaluated using [¹¹C]PiB PET, whereas tau accumulation was assessed using florzolotau (18F) PET. PET scans were obtained using Siemens or GE scanners according to standardized protocols. Florzolotau (18F) PET images were obtained 90–110 min after tracer injection, and standardized uptake value ratio (SUVR) images were generated using a data driven gray matter reference region.

Detailed PET acquisition and reconstruction procedures are described in the eMethods: *PET Acquisition and Image Reconstruction, and Radioligand Synthesis* section.

[^11^C]PiB and florzolotau (18F) PET scans were completed within a 3-month interval. The PET scan protocol in detail was provided elsewhere.^11,29,30^

### Data and Statistical Analysis

Voxel-based analysis was conducted using tau PET images transformed into the standard brain space via DARTEL (Details are provided in the eMethods; *Data Preprocessing* for details). Tau accumulation and brain atrophy in PSP versus HC subjects were analyzed by identifying regions of voxels with a family-wise error correction at peak-level (FWEp) of *p* < 0.05 and the voxel number exceeding assigned values. The association of tau accumulation and brain volume with neurological symptoms was evaluated to pick up voxel clusters with predefined height and extent thresholds. The association of tau depositions and brain volume with neurological symptoms was assessed using a threshold of FWE correction at cluster-level (FWEc) *p* < 0.05. All PSP-versus-HC comparisons and all analyses examining associations between imaging measures and clinical scores within the PSP cohort were adjusted for age and sex. Disease duration was not included as a covariate because no significant voxel-wise associations were observed between florzolotau (18F) uptake and disease duration.

Based on voxel-wise analyses, areas of tau accumulation and volume loss associated with each neurological symptom were binarized as ROIs. ROIs included all significant clusters identified from tau PET and MRI analyses without restricting inclusion to overlapping or modality-specific voxels (i.e., tau-only or atrophy-only regions). Using these ROIs, florzolotau (18F) SUVRs and brain volumes were calculated for each PSP case.

For path analysis, PSP Rating Scale subdomain scores, ROI-based florzolotau (18F) SUVRs, and regional brain volumes were residualized for age and sex. A single-mediator path model (mediation framework) was specified in which tau deposition, represented by florzolotau (18F) SUVRs, served as the predictor, regional atrophy as the mediator, and domain-specific symptom severity as the outcome. Effects were evaluated as hypothesis-driven associations consistent with this biologically motivated causal ordering. Details of the analytical workflow are provided in the supplementary flowchart (eSAP 2).

To investigate whether these hypothesized associations varied across disease stages, additional path analyses were conducted after stratifying participants according to disease duration. Patients with PSP were categorized into three groups based on disease duration (1 year, 2 years, and ≥3 years), and identical path models were applied within each subgroup. These disease-duration-stratified analyses correspond to the results presented in Figure 3.

Path analyses were performed to evaluate hypothesized relationships among tau deposition, regional atrophy, and domain-specific symptom severity. Detailed procedures are described in the eMethods: *Statistical and Path Analysis Procedures* section.

### Data Sharing and Data Availability

Requests for data supporting the findings of this study should be directed to the corresponding author, Hironobu Endo. The data supporting our study findings are available from the corresponding author upon reasonable request.

### Standard Protocol Approvals, Registrations, and Patient Consents

The study protocol was approved by the Ethics Committee of the National Institutes for Quantum Science and Technology. Written informed consent was obtained from all participants. The study was registered in the UMIN Clinical Trials Registry (registration numbers 000026385, 000029608, and 000030248).

## Results

### Patient Backgrounds

The study included 52 HCs and 58 patients with PSP. The PSP cohort had a mean age of 71.6 ± 7.7 years, 43.1% were women, and the mean disease duration was 3.9 ± 3.2 years. The mean PSP Rating Scale total score was 36.2 ± 16.5. Patients with PSP were older than HCs, whereas sex distribution did not significantly differ between groups. As expected, patients with PSP exhibited substantially higher PSP Rating Scale scores, indicating greater neurological impairment. The demographic profiles of the PSP patients and HCs are provided in Table 1.

**Table 1:** Demographic characteristics.

|  | HC (n = 52) | PSP (n = 58) | P-value |
| --- | --- | --- | --- |
| Women, n (%) | 24 (45.3) | 25 (43.1) | 0.85 |
| Age, mean ± SD (years) | 60.8 ± 13.3 | 71.6 ± 7.7 | < 0.001 |
| Disease duration, mean ± SD (years) | N/A | 3.9 ± 3.2 | N/A |
| PSP Rating Scale total (point),<br>mean ± SD/(maximum) | 0.6 ± 1.3/100 | 36.2 ± 16.5/100 | < 0.001 |
| History (point),<br>mean ± SD/(maximum) | 0.0 ± 0.1/24 | 8.7 ± 4.7/24 | < 0.001 |
| Mentation (point),<br>mean ± SD/(maximum) | 0.0 ± 0.0/16 | 2.7 ± 3.1/16 | < 0.001 |
| Bulbar (point),<br>mean ± SD/(maximum) | 0.0 ± 0.0/8 | 2.8 ± 1.9/8 | < 0.001 |
| Ocular motor (point),<br>mean ± SD/(maximum) | 0.1 ± 0.5/16 | 7.2 ± 3.8/16 | < 0.001 |
| Limb motor (point),<br>mean ± SD/(maximum) | 0.4 ± 0.9/16 | 4.3 ± 2.3/16 | < 0.001 |
| Gait and midline (point),<br>mean ± SD/(maximum) | 0.0 ± 0.2/20 | 10.5 ± 5.0/20 | < 0.001 |
| FAB/(maximum) | 16.6 ± 1.1/18 | 12.0 ± 4.1/18 | <0.001 |

Data are expressed as the number (%), mean ± SD for normally distributed variables.
HC, healthy control; PSP, progressive supranuclear palsy; FAB, Frontal

### Exhaustive Voxel-based Assessments

#### Comparisons between the PSP and HC groups

The retention of florzolotau (18F) was enhanced in the subcortical structures, such as the midbrain, basal ganglia, and frontal regions in PSP cases relative to HCs (red and yellow in Figure 1), in line with the distribution of tau pathologies in this illness. These patients also presented reduced volumes of characteristic brain areas, including the midbrain, basal ganglia, thalamus, and cerebellar hemispheres, compared with HCs (green and yellow in Figure 1). There were notable spatial overlaps between tau depositions and atrophy in the midbrain and basal ganglia (yellow in Figure 1). Representative florzolotau (18F) PET images are shown in Figure 1B, illustrating typical tracer uptake patterns in an HC, a patient with Richardson’s syndrome (RS)-PSP, and a patient with non RS PSP (PSP-parkinsonism [PSP P]).

**Figure 1:**
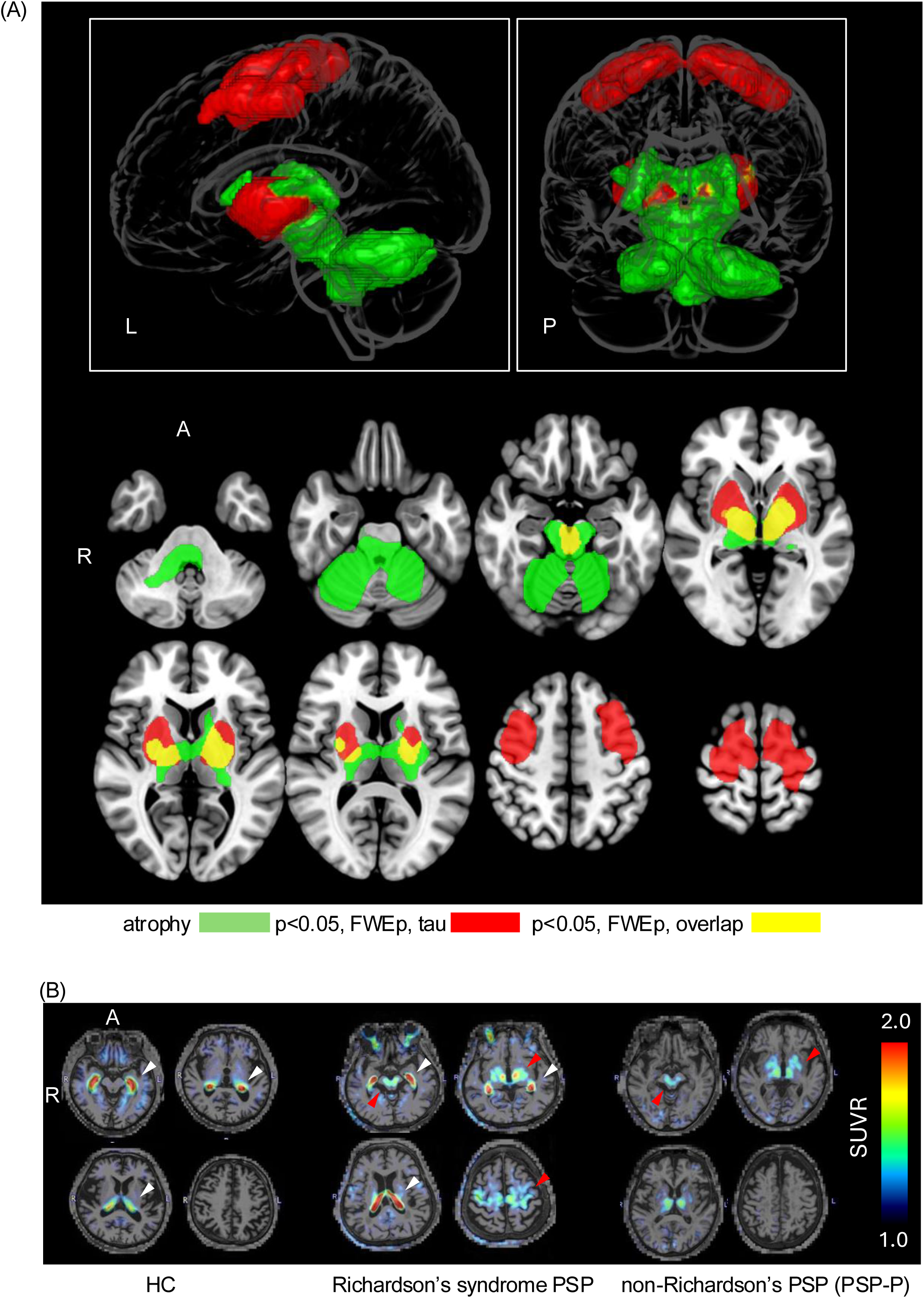
Topologies of tau accumulation and atrophy in the brains of PSP patients relative to controls. Voxel-based analysis showed an elevated tau PET tracer retention (red) primarily in the basal ganglia and frontal lobes, atrophy (green) in the basal ganglia, thalamus, and cerebellar hemispheres, and their spatial overlaps (yellow) in the midbrain and basal ganglia of the PSP group relative to HCs (*p* < 0.05, FWEp). Cluster size thresholds (k) are 115 voxels for tau accumulation and 91 voxels for atrophy. Areas are highlighted in the standard anatomical space of the brain. (B) Representative raw florzolotau (18F) PET images are shown for an HC (left), a patient with Richardson’s syndrome PSP (middle), and a patient with non Richardson’s PSP (PSP P) (right). White arrowheads indicate the choroid plexus, a known site of off-target tracer binding. In PSP-Richardson’s syndrome, tracer retention was observed not only in the subthalamic nucleus, adjacent thalamic and basal ganglia regions, and the midbrain, but also extended into the neocortex, including the primary motor cortex and adjacent white matter (red arrowheads). In contrast, in PSP-P, tracer retention was largely confined to the subthalamic nucleus, adjacent thalamic and basal ganglia regions, and the midbrain, with lower intensity than that observed in PSP-Richardson’s syndrome. PSP, progressive supranuclear palsy; HC, healthy control; FWEp, family-wise error correction at the peak level; PSP P, PSP-parkinsonism.

#### Topology of tau accumulation and volume loss associated with neurological symptoms

##### (A) Total PSP Rating Scale scores

Intensified florzolotau (18F) signals and concomitant volume declines in subcortical locations, including the midbrain, subthalamic nucleus, and globus pallidus, were associated with higher total PSP Rating Scale scores (upper left panels in Figure 2). In contrast, tau accumulation in neocortical regions such as the primary and supplementary motor cortices, was also associated with symptom severity despite the absence of marked local atrophy.

**Figure 2:**
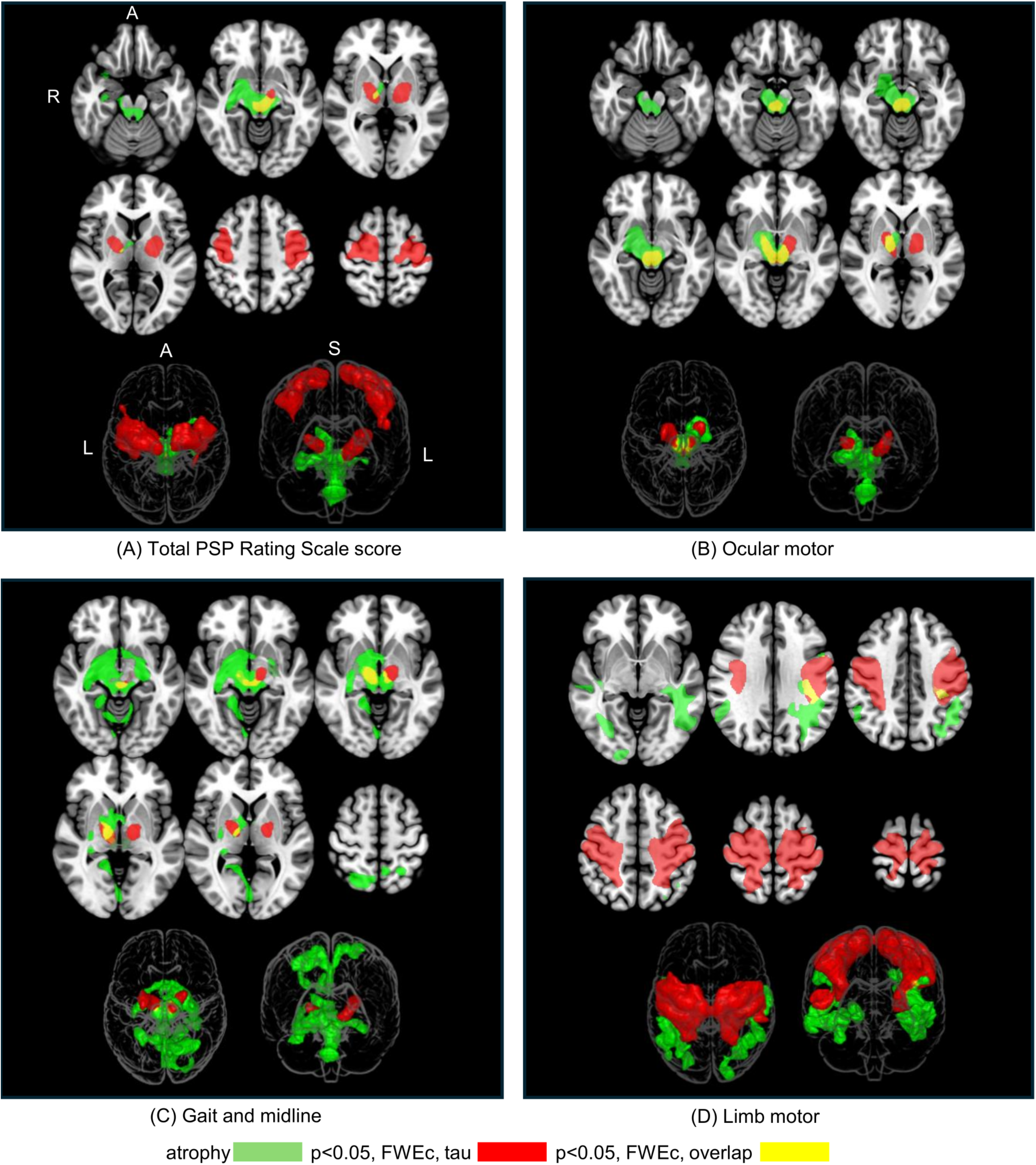
Distributions of tau accumulation (red), atrophy (green), and their overlaps (yellow) associated with neurological symptoms. (A) Total PSP Rating Scale scores were associated with both tau accumulation and atrophy in the subthalamic nucleus and globus pallidus. Associations with tau accumulation was also found in neocortical regions, including the primary and supplementary motor cortices. Cluster size thresholds (k) are 4229 voxels for tau accumulation and 7338 voxels for atrophy. (B) “Ocular motor” subscores were associated with tau accumulation and atrophy commonly in the midbrain tegmentum and globus pallidus. Cluster size thresholds (k) are 3658 voxels for tau accumulation and 7031 voxels for atrophy. (C) “Gait and midline” subscores were associated with tau accumulation in the globus pallidus to thalamic area and occipital cortex and atrophy in the globus pallidus to the thalamic area, occipital cortex, and precuneus. Cluster size thresholds (k) are 2480 voxels for tau accumulation and 2326 voxels for atrophy. (D) “Limb motor” subscores were associated with tau accumulation in the primary motor and somatosensory cortices and atrophy in the angular and supramarginal gyri and temporal gray and white matter. Cluster size thresholds (k) are 43882 voxels for tau accumulation and 2489 voxels for atrophy. Areas are highlighted in the standard anatomical space of the brain. All statistical thresholds were set at *p* < 0.05, FWEc. PSP, progressive supranuclear palsy; FWEc, family-wise error correction at the cluster-level.

##### (B) Ocular motor symptom domain

Tau accumulations and reduced volumes in subcortical structures, including the midbrain tegmentum and globus pallidus, were associated with elevated PSP Rating Scale subscores in the ocular motor domain (upper right panels in Figure 2). The voxels highlighted in PET and MRI assays presented remarkable spatial overlaps.

##### (C) Gait and midline symptom domain

Tau depositions and volume reductions in subcortical structures, including the subthalamic nucleus and surrounding thalamic regions, alongside the globus pallidus, were associated with increases of PSP Rating Scale subscores in the gait and midline domain (lower left panels in Figure 2). The voxels highlighted in tau PET and MRI assays displayed notable overlaps in these locations. In addition, we observed associations of tau accumulations in the occipital cortex and volume loss in the occipital cortex and precuneus with this symptomatic domain. The neocortical voxels highlighted by tau PET and MRI showed little spatial overlap.

##### (D) Limb motor symptom domain

Unlike the symptom domains (A)-(C), only limited spatial overlap was observed between regions in which tau deposition and volume loss were associated with limb motor impairment (lower right panels in Figure 2). Tau accumulation, as related to the PSP Rating Scale limb motor subscores, mainly involved the primary motor and somatosensory cortices. In contrast, symptom-related atrophy was predominantly localized to the angular and supramarginal gyri and temporal lobes, including both gray and white matter. Although minor overlap near boundaries was present, the principal foci of tau lesions and atrophy were spatially dissociated.

Three-dimensional renderings of all tau-loaded and atrophic regions associated with domains (A)-(D) are shown in eFigures 1 and 2.

To facilitate interpretation of these findings at the individual level for symptom domains (B)–(D), associations among ROI-based tau burden, regional brain volume, and domain-specific PSP Rating Scale subscores are shown in the Supplementary Material (eFigure 3).

##### (E) Other categories

We found no brain areas exhibiting associations of tau accumulations and atrophy with PSP Rating Scale subscores in history, mentation, and bulbar categories.

### Path Analysis Exploring Hypothesized Directionality

Areas of tau accumulation and volume loss associated with each neurological symptom were binarized as ROIs. Using these ROIs, florzolotau (18F) SUVRs and brain volumes were calculated for each PSP case.

Path analysis was performed using ROI values and PSP Rating Scale subscores to evaluate the hypothesized relationships among tau deposition, regional brain volume, and symptom severity in the cross-sectional data (eFigure 4).

In analyses including all patients, regional brain volume demonstrated a stronger association with symptom severity than tau burden (eFigure 4). This pattern may reflect stronger structure–symptom relationships in patients with longer disease duration. Therefore, patients were stratified into early-, intermediate-, and advanced-stage groups based on disease duration, and path analyses were repeated within each subgroup (Fig. 3 and eFigure 5). The demographic and clinical profiles of patients with PSP in each disease-duration group are summarized in Table 2 and illustrated in eFigure 6. In the ocular motor domain, the association between tau burden and regional atrophy was more prominent in the early and intermediate disease-duration groups, whereas the association between regional atrophy and symptom severity was stronger in the advanced group.

**Figure 3:**
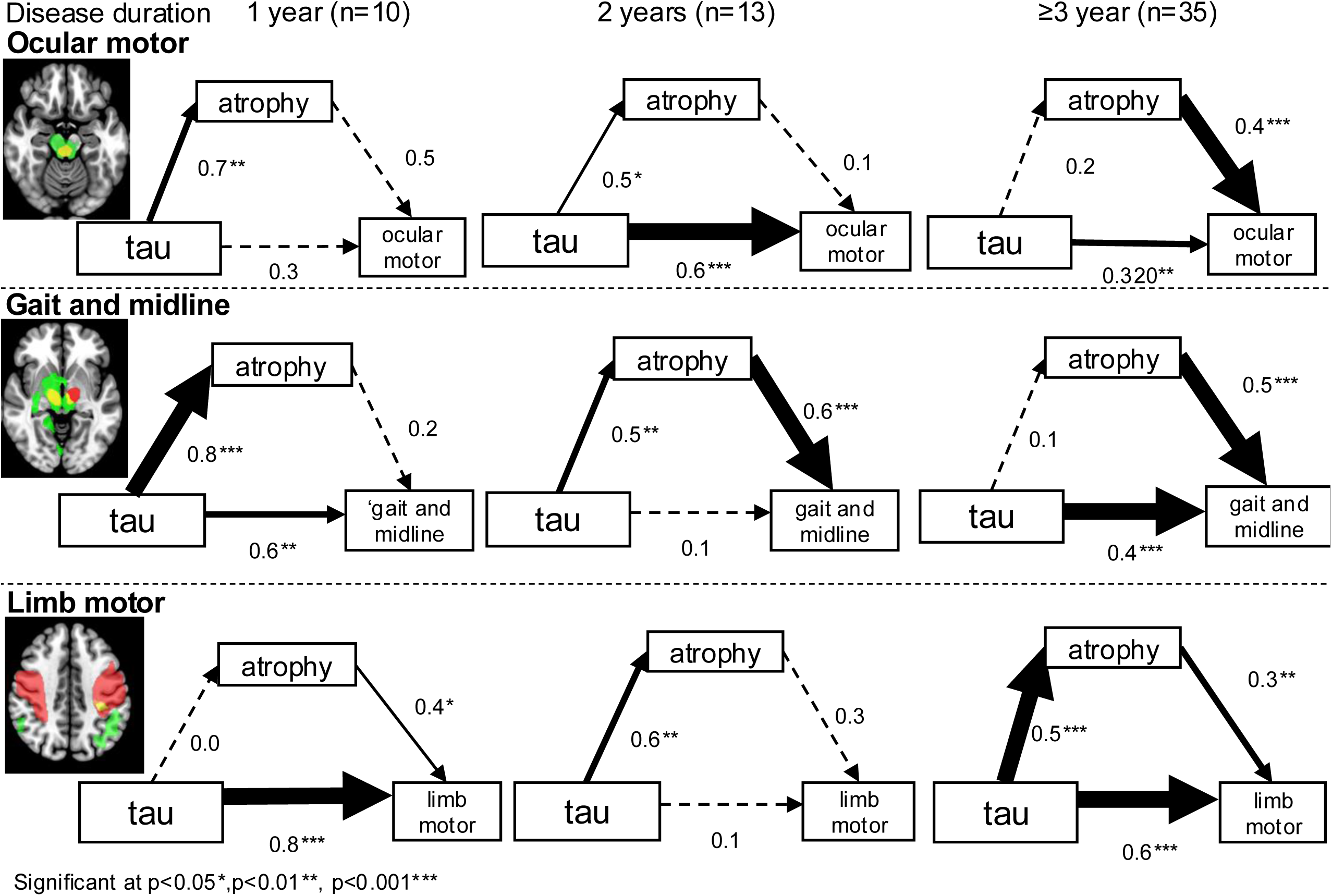
Path analysis of associations among tau deposition, brain atrophy, and clinical symptoms across disease-duration strata. Path analyses were conducted across three disease-duration groups (early, 1 year; intermediate, 2 years; advanced, ≥3 years) for the ocular motor, gait and midline, and limb motor symptom domains. Each panel illustrates the associations among tau deposition measured by florzolotau SUVR, regional brain volume, and domain-specific symptom severity assessed using the PSP Rating Scale. Numbers represent the absolute values of standardized regression coefficients (β), with thicker arrows indicating larger coefficients. Tau PET is expressed as florzolotau SUVR, and volume represents regional brain volume. All analyses were performed using values residualized for age and sex. Asterisks indicate statistical significance (\**p* < 0.05, \*\**p* < 0.01, \*\*\**p* < 0.001).

**Table 2:** Demographic characteristics in three disease-duration subgroups.

|  | Disease duration |  |  | P-value |
| --- | --- | --- | --- | --- |
|  | 1 year (n = 10) | 2 years (n = 13) | ≥ 3 years (n = 35) |  |
| Women, n (%) | 2 (20.0) | 6 (46.2) | 17 (48.6) | 0.265 |
| Age (years) | 68.2 ± 8.6 | 74.1 ± 8.4 | 71.7 ± 7.0 | 0.192 |
| Disease duration (years) | 1.0 ± 0.0 | 2.0 ± 0.0<sup>####</sup> | 5.4 ± 3.4<sup>****</sup> | <0.001 |
| PSP Rating Scale total(point) | 27.2 ± 10.9/100 | 26.5 ± 11.6<sup>##</sup>/100 | 42.4 ± 16.7<sup>*</sup>/100 | 0.002 |
| History (point) | 6.4 ± 4.0/24 | 5.6 ± 3.4<sup>##</sup>/24 | 10.4 ± 4.6/24 | 0.005 |
| Mentation (point) | 1.8 ± 1.8/16 | 1.9 ± 2.1/16 | 3.3 ± 3.6/16 | 0.465 |
| Bulbar (point) | 2.0 ± 1.5/8 | 1.3 ± 0.8<sup>###</sup>/8 | 3.6 ± 1.9/8 | <0.001 |
| Ocular motor (point) | 6.6 ± 3.1/16 | 5.6 ± 3.0/16 | 7.9 ± 4.1/16 | 0.15 |
| Limb motor (point) | 3.3 ± 1.6/16 | 4.1 ± 1.7/16 | 4.7 ± 2.6/16 | 0.324 |
| Gait and midline (point) | 7.1 ± 4.2/20 | 8.0 ± 5.2<sup>#</sup>/20 | 12.4 ± 4.2<sup>**</sup>/20 | 0.002 |
| FAB (point) | 12.3 ± 4.3/18 | 12.8 ± 3.0/18 | 11.6 ± 4.2/18 | 0.64 |

Data are expressed as the number (%) or mean ± SD, as appropriate; age, disease duration, PSP Rating Scale total and subdomain scores, and FAB score are expressed as mean ± SD.
\*\*\*\*P < 0.0001, \*\*P < 0.01, \*P < 0.05 versus 1 year group post hoc Dunn's test

In the gait and midline domain, the association between tau burden and regional atrophy was evident in the early-stage group, whereas the association between regional atrophy and symptom severity emerged in the intermediate-stage group. In the limb motor domain, tau burden exhibited a stronger association with symptom severity in the early-stage group. In the later-stage groups, the model incorporated tau burden, atrophy in spatially distinct ROIs, and symptom severity.

Results restricted to patients with PSP-RS and their clinical characteristics are presented in eFigures 7–9 and eTable 1, with corresponding interpretations provided in eAppendixes 1 and 2. PSP subtype-specific patterns of tau accumulation and atrophy, together with the clinical characteristics of each subtype, are presented in eFigure 10 and eTable2, with detailed interpretations described in eAppendix 3. Furthermore, voxel-wise analyses using residualized domain scores adjusted for total PSP Rating Scale score were performed to evaluate domain-specific associations with tau accumulation and atrophy (eFigure 11 and eAppendix 4).

## Discussion

In this study, we generated high-contrast maps delineating tau pathology and regional volume loss in the brains of PSP cases. Our analyses revealed, for the first time, the spatial distribution of tau deposits and brain atrophy in relation to core symptomatic domains captured by the PSP Rating Scale. Clinically, these domain-anchored topographies may facilitate the linkage of symptom profiles to their underlying neuroanatomical substrates and may inform diagnostic interpretation as well as biomarker development. Such approaches may ultimately support the earlier detection of tau pathology before structural degeneration becomes apparent. Integrating these findings with path analyses, our results suggest that subcortical tau accumulation is associated with local atrophy and symptom severity, whereas neocortical tau deposition may be associated with more distributed patterns of neurodegeneration. These findings support distinct mechanisms of subcortical and neocortical involvement in PSP.

Comparisons between the PSP and control groups revealed a spatial concordance of tau pathology and volume reduction. In contrast, similarly prominent neocortical volume reductions were not observed alongside neocortical tau retention. These findings suggest that morphometric alterations in PSP are predominantly subcortical, while neocortical neuronal loss may be more heterogeneous across individuals. Indeed, reduced volumes in posterior cortical areas were evident in patients exhibiting severe “gait and midline” and “limb motor” symptoms (Fig. 2C, D).

The relative preservation of primary motor cortex volume despite tau accumulation has been reported previously and may reflect region-specific vulnerability (details are provided in eAppendix 5).^15^ We observed substantial spatial overlap between regions of tau accumulation and brain atrophy, which were closely associated with total PSP Rating Scale scores and subscores in the “ocular motor” and “gait and midline” domains. These voxel-wise analyses primarily identified subcortical structures. Notably, impairments in the “ocular motor” domain showed strong associations with both tau pathology and volume loss in the midbrain.

Our tau and atrophy maps highlighted medial midbrain regions encompassing the rostral interstitial nucleus of the medial longitudinal fasciculus (riMLF), consistent with its established role in ocular motor dysfunction.^31^ Importantly, the superior colliculus (SC) plays a modulatory role in the activity of riMLF, which is critical for controlling eye movements.^32^ Disruptions in the sequential pathways linking the globus pallidus to the SC—via the subthalamic nucleus and substantia nigra pars reticulata (SNr)—have been implicated in the pathophysiology of ocular motor dysfunction.^33,34^ Consistently, our analysis identified the globus pallidus, subthalamic nucleus, and SNr as regions showing significant associations between tau deposition, structural atrophy, and impairment in the “ocular motor” domain. Furthermore, exploratory path analysis (upper panel of Fig. 3) indicated that the cross-sectional associations among tau burden, regional atrophy, and ocular motor symptom severity were statistically consistent with an ordered relationship across disease-duration strata. These findings align with previous clinical reports showing that gaze abnormalities are increasingly prevalent during later stages of PSP, particularly among nonclassical phenotypes.^35^

As mentioned above, regions exhibiting tau deposition and volume loss associated with the “gait and midline” domain substantially overlapped with subcortical areas implicated in “ocular motor” deficits, whereas the brainstem appeared to be less involved. The subcortical locomotive network includes the striatum, globus pallidus, subthalamic nucleus, mesencephalic locomotor region, and pontine and medullary reticular formations.^36,37^ Among these, the subthalamic nucleus and basal ganglia (highlighted yellow in Fig. 2B and the middle-left panel of Fig. 3) were prominently identified in our analyses. These structures are implicated from the early stages of PSP, including phenotypes with spatially restricted tau accumulation such as PSP-P and PSP-pure akinesia with gait freezing (PSP-PGF),^38^ and are accordingly more consistently involved in locomotive impairment than the brainstem.

Gait disturbances, particularly balance difficulties and falls, frequently represent the earliest clinical manifestations of PSP, reinforcing the association between these subcortical structures and the “gait and midline” subscores. Correspondingly, path analysis (middle panels of Fig. 3) suggested a hypothesized pathway from early-stage tau accumulation to impairments in this functional domain within one year of symptom onset. In addition, neocortical involvement may contribute to “gait and midline” symptoms through distributed network effects (details are provided in eAppendix 6).

Unlike the “ocular motor” and “gait and midline” domains, subscores of the “limb motor” domain was primarily associated with pathological changes in the neocortex. Regions of tau deposition and atrophy associated with this domain were spatially distinct (Fig. 2D).

We found that volume loss in the angular and supramarginal gyri and temporal lobes was strongly associated with limb motor deficits, consistent with the recognized role of parietal regions to limb apraxia.^2,39^ Tau accumulation within the primary motor and somatosensory cortices may exert secondary effects on downstream regions. The superior longitudinal fasciculus, which connects the primary motor cortex and angular gyrus, may provide a pathway for transneuronal degeneration, thereby contributing to volume loss in these regions^40^ (see eAppendix 7 for details).

Although the domain also includes Parkinsonian features such as limb rigidity and tremor, subcortical regions typically associated with the extrapyramidal system were not prominently implicated in the present analysis. This may reflect the item composition of the domain, which emphasizes functions, such as dystonia and fine motor tasks (e.g., finger and toe tapping), that are more reliant on cortical motor circuits.^41^ ^42^

Tau deposition in the supplementary and primary motor cortices was associated with the mentation domain of the PSP Rating Scale, whereas involvement of the supplementary and pre-supplementary motor cortices and the inferior frontal cortex (IFC) was associated with FAB scores (eFigure 12), indicating that tau pathology in these regions contributes to frontal lobe function.^43^ Notably, FAB scores were not associated with regional atrophy. In contrast, our recent work restricted analysis to patients with PSP-RS identified a ‘latent’ network containing the dorsomedial frontal cortex (dmFC), premotor cortex (PMC), and IFC associated with FAB scores in the absence of local tau accumulations.^44^ In the current study, however, FAB scores were associated with tau deposition in cortical areas corresponding to the dmFC, PMC, and IFC. The discrepancy between studies may reflect differing cohort compositions; our cohort included both PSP-RS (n = 43) and non-RS subtypes (n = 15; PSP-P, n = 10; PSP with frontal cognitive or behavioral presentations, n = 4; PSP-PGF, n = 1).

Collectively, these findings suggest that the association between tau pathology and executive dysfunction, particularly within the frontal lobe, varies depending on the PSP subtypes included in the analysis.

Potential methodological and biological explanations for the absence of significant imaging associations in the history, mentation, and bulbar domains are discussed in eAppendix 8.

Our path analysis (lower panel of Fig. 3) suggested a directional relationship from early-stage tau deposition to limb motor impairments by less than one year of onset, indicating that tau-induced dysfunction within cortical motor areas may precede significant neuronal loss. As the disease progresses, atrophy in distant projection sites likely emerges, contributing to the focal and network-level motor deficits observed in advanced PSP.

Strengths of this study include the use of high-contrast florzolotau (18F) PET imaging in a relatively large, well-characterized PSP cohort, enabling detailed voxel-wise and domain-specific analyses. Anchoring molecular and structural imaging findings to PSP Rating Scale subdomains provides clinically interpretable topographies linking tau pathology to regional brain involvement, supported by complementary ROI-based and exploratory path analyses.

Despite these findings demonstrating localized associations between tau pathology, neuronal loss, and symptom domains, the broader impact of tau deposition on network-level degeneration, including remote atrophy and functional impairment, remains unclear and warrants further investigation using tract-specific approaches such as diffusion tensor imaging. In addition, several limitations should be acknowledged. Although analyses were performed in PSP-RS, pathology–symptom relationships were not fully stratified across PSP subtypes. A further limitation is potential circularity between ROI definition and path modeling, as ROIs derived from voxel-wise symptom associations within the same dataset were reused in path analyses; these findings should therefore be considered exploratory and require independent validation. While path analysis suggested temporal relationships linking tau accumulation and regional atrophy to specific symptom domains, longitudinal neuroimaging studies are required for confirmation. Phenotypic stratification was limited by incomplete assessment of standardized levodopa responsiveness, although all patients were classified by specialists using Movement Disorder Society diagnostic criteria, ensuring high diagnostic certainty. Finally, as a single-center cross-sectional study, causal and temporal inferences are limited. Although analyses were adjusted for age and sex, residual confounding may persist, and incomplete age matching between patients and controls may affect group comparability.

In conclusion, by leveraging high-contrast PET imaging of four-repeat tau assemblies, the present study delineates the spatial patterns of neurodegenerative tau pathologies underlying the emergence and progression of specific symptom domains in PSP. This approach holds promise for predicting clinical outcomes across the natural course of the disease and in the context of anti-tau therapeutic interventions.

## Supporting information

eFigure

eSAP

eMethods

eAppendix

eTable

eReference

## Data Availability

All data produced in the present study are available upon reasonable request to the authors

## Acknowledgment

This study was supported by AMED under grants JP19dm0207072, JP24wm0625001, JP24zf0127012, JP24wm0625307; MEXT KAKENHI grant JP21K15705; JST CREST grants JPMJMS2024; Biogen Inc.; and APRINOIA Therapeutics. We thank all patients and their caregivers, as well as volunteers, for their participation in this study; clinical research coordinators; PET and MRI operators; radiochemists; and research ethics advisers at Quantum Science and Technology (QST) for their assistance with the current projects. We acknowledge the support of M. Kubota, Y. Yamamoto, and S. Kitamura at QST. We thank APRINOIA Therapeutics for kindly sharing the precursor of florzolotau (18F). We acknowledge the support of S. Hirano at the Department of Neurology, Chiba University on patient recruitment; T. Hatano, T. Tsunemi, N. Nishikawa, K. Nishioka (currently working at The Juntendo Tokyo Koto Geriatric Medical Center), Y. Yamashita, Y. Motoi, and S. Saiki (currently working at the University of Tsukuba) at the Department of Neurology, Juntendo University School of Medicine; I. Aiba at the Department of Neurology, National Hospital Organization Higashinagoya National Hospital; Y. Nakano (currently working at The Feinstein Institute for Medical Research) at the Department of Neurology, Chibaken Saiseikai Narashino Hospital; T. Yuasa at the Department of Neurology, Kamagaya General Hospital; H. Imai at the Tokyo Rinkai Hospital; Y. Nishida and Y. Yagi at the Department of Neurology, Tokyo Medical and Dental University; S. Furukawa at the Narita Red Cross Hospital; M. Seki at the Department of Neurology, Keio University School of Medicine; and T. Takeda and I. Isose at the Department of Neurology, Chiba-East Hospital.

## Disclosure

Y. Kataoka reports no disclosures. R. Goto reports no disclosures. Y. Chishiki reports no disclosures. K. Osawa reports no disclosures. A. Oyama reports no disclosures. H. Matsumoto reports no disclosures. M. Ichihashi reports no disclosures. S. Moriguchi reports no disclosures. Y. Momota reports no disclosures. T. Kamada reports no disclosures. C. Seki reports no disclosures. K. Matsuoka reports no disclosures. K. Hirata reports no disclosures. S. Kurose reports no disclosures. M. Oya reports no disclosures. N. Kokubo reports no disclosures. H. Shinotoh reports no disclosures. H. Shimada, M.-R. Zhang, and M. Higuchi hold patents related to compounds described in this report (JP 5422782/EP 12 884 742.3/CA2894994/HK1208672/ZL201710407246.4). K. Kawamura reports no disclosures. T. Tokuda reports no disclosures. K. Takahata reports no disclosures. K. Tagai reports no disclosures. H. Endo reports no disclosures.

## Data Access

The Principal Author had full access to all the data used in this study and takes full responsibility for the integrity of the data, the accuracy of the analyses, and the decision to publish the results.

## Author Contributions

Y.K., M.H., and H.E. contributed to drafting and revision of the manuscript for content, including medical writing; Y.K., R.G., Y.C., K.O., H.M., M.I., S.M., Y.M., T.K., K.M., K.H., S.K., M.O., K. Takahata, and N.K. contributed to the acquisition of data; Y.K., M.H., H.E., H. Shinotoh, H. Shimada, and K. Tagai contributed to the study concept and design; Y.K., A.O., C.S., K.K., M.-R.Z., T.T., M.H., and H.E. contributed to the analysis and interpretation of data.

## Notes

### Competing Interest Statement

H. Shimada, M.-R.Z., and M.H. hold patents on compounds related to this report (JP 5422782/EP 12 884 742.3/CA2894994/HK1208672/ZL201710407246.4).

### Author Declarations

Standard Protocol Approvals, Registrations, and Patient Consents The study protocol was approved by the Ethics Committee of the National Institutes for Quantum Science and Technology. Written informed consent was obtained from all participants. The study was registered in the UMIN Clinical Trials Registry (registration numbers 000026385, 000029608, and 000030248).

### Summary of Updates

This version has been revised to address reviewer comments. The manuscript text has been clarified, additional analyses and supplemental materials have been added, figures have been updated, and minor editorial corrections have been made.

