## Supplementary material for "*In Vivo* Topographic Associations Between Tau Pathology, Atrophy, and Symptom Domains in Patients with Progressive Supranuclear Palsy": eFigure

**Supplementary materials**

Supplementary material associated with this study can be found online.

Abbreviations: HC, healthy controls; PSP, progressive supranuclear palsy

**eFigures**

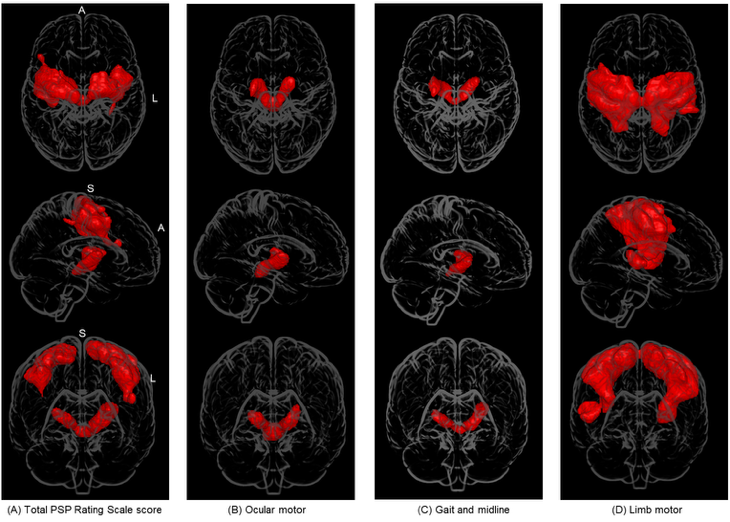
**eFigure 1: Topologies of tau accumulation associated with neurological symptoms in the 3-D standard anatomical space of the brain**

Voxel clusters showing significant association between tau probe retention and total scores of PSP Rating Scale (A) (FWEc *p <* 0.05, k = 4229 voxels) and subscores of “ocular motor” (B) (FWEc *p <* 0.05, k = 3658 voxels), “gait and midline” (C) (FWEc *p <* 0.05, k = 2480 voxels), and “limb motor” (D) (FWEc *p <* 0.05, k = 43882 voxels) symptomatic domains are highlighted in the 3-D glass brain. Subcortical structures are predominantly picked up in the analyses (B) and (C), while clusters are primarily localized to neocortical areas in the analysis (D). Both subcortical and neocortical regions are involved in the analysis (A).

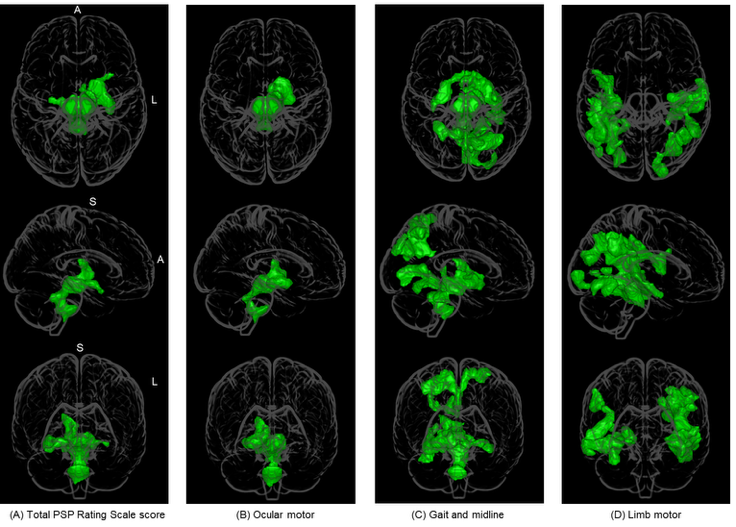
**eFigure 2:** **Topologies of atrophy associated with neurological symptoms in the 3-D standard anatomical space of the brain**

Voxel clusters showing significant association between volume reduction and total scores of PSP Rating Scale (A) (FWEc *p <* 0.05, k = 7338 voxels) and subscores of “ocular motor” (B) (FWEc *p <* 0.05, k = 7031 voxels), “gait and midline” (C) (FWEc *p <* 0.05, k = 2326 voxels), and “limb motor” (D) (FWEc *p <* 0.05, k = 2489 voxels) symptomatic domains are highlighted in the 3-D glass brain. Subcortical structures are predominantly picked up in the analyses (A) - (C), whereas clusters are primarily localized to neocortical areas in the analysis (D).

**eFigure 3: Relationships between tau accumulation and brain atrophy in symptom‑related ROIs and PSP Rating Scale scores**

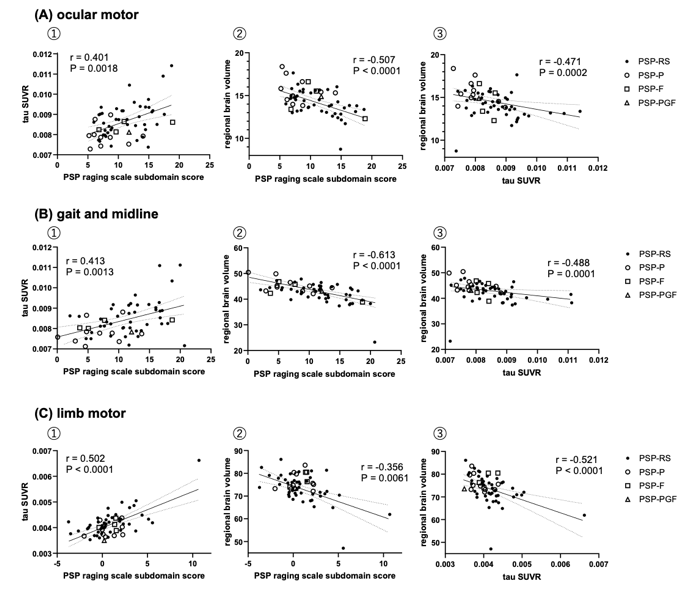

Scatterplots illustrate the relationships between neuroimaging measures within regions of interest (ROIs) associated with specific neurological symptom domains and PSP Rating Scale subscores. The solid line represents the linear regression fit, and the dashed lines indicate the 95% confidence interval. Spearman’s correlation coefficient (r) and *p*-value are displayed in each panel.

The symptom domains analyzed were (a) ocular motor, (b) gait and midline, and (c) limb motor. For each domain, tau PET metrics (florzolotau (18F) standardized uptake value ratio [SUVR]) and regional brain volumes (mL) were extracted from ROIs identified as being associated with the corresponding symptom domain.

Panel ① shows the relationship between PSP Rating Scale subscores and tau SUVR, panel ② shows subtype-specific relationships between PSP Rating Scale subscores and tau SUVR, and panel ③ shows the relationship between tau SUVR and regional brain volume. All analyses were performed using age- and sex-adjusted values. These scatterplots were generated to facilitate visual assessment of the strength and linearity of the associations and to identify potential outliers. Tau SUVR values represent the mean SUVR across all voxels within each ROI.

The number of voxels included in the ROIs was 3,658 for the ocular motor domain, 2,480 for the gait and midline domain, and 43,882 for the limb motor domain.

RS, Richardson’s syndrome; P, parkinsonism; F, frontal lobe cognitive or behavioral presentations; PGF, pure akinesia with gait freezing.

**eFigure 4: Path analysis among tau probe retention, brain atrophy, and neurological symptoms in all patients with PSP**

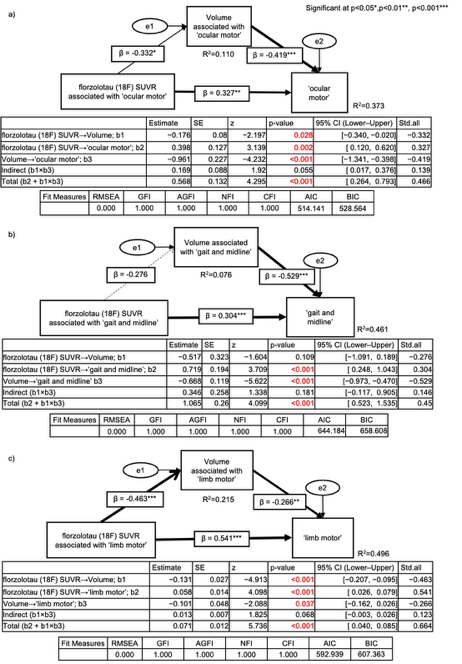

Path analysis was performed to examine hypothesized associations among tau probe SUVR, regional brain volume, and symptom severity in the “ocular motor” (a), “gait and midline” (b), and “limb motor” domains. Neurological symptom severity was associated with both regional brain volume and tau probe SUVR, with stronger associations observed for regional brain volume.

SUVR, standardized uptake value ratio; SE, standard error; CI, confidence interval; Std. all, fully standardized coefficients; RMSEA, Root Mean Square Error of Approximation; GFI, Goodness‑of‑Fit Index; AGFI, Adjusted Goodness‑of‑Fit Index; NFI, Normed Fit Index; CFI, Comparative Fit Index; AIC, Akaike Information Criterion; BIC, Bayesian Information Criterion.

**eFigure 5: Path analysis among tau probe retention, brain atrophy, and neurological symptoms in groups classified by disease duration**
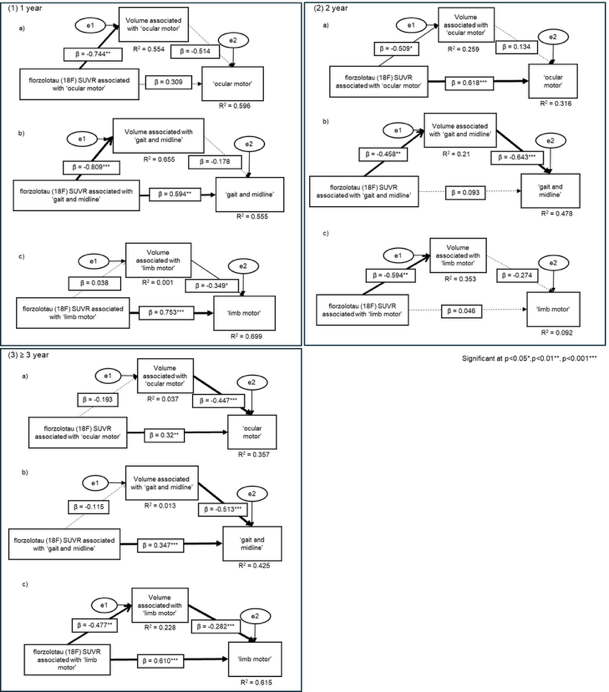

Path analysis was performed to examine hypothesized associations among tau probe SUVR, regional brain volume, and symptom severity for the “ocular motor” (a), “gait and midline” (b), and “limb motor” (c) domains in patients with disease duration of 1 year (1), 2 years (2), and ≥3 years (3).

SUVR, standardized uptake value ratio.

**eFigure 6: Distribution of disease duration and overall disease severity in the PSP cases.**

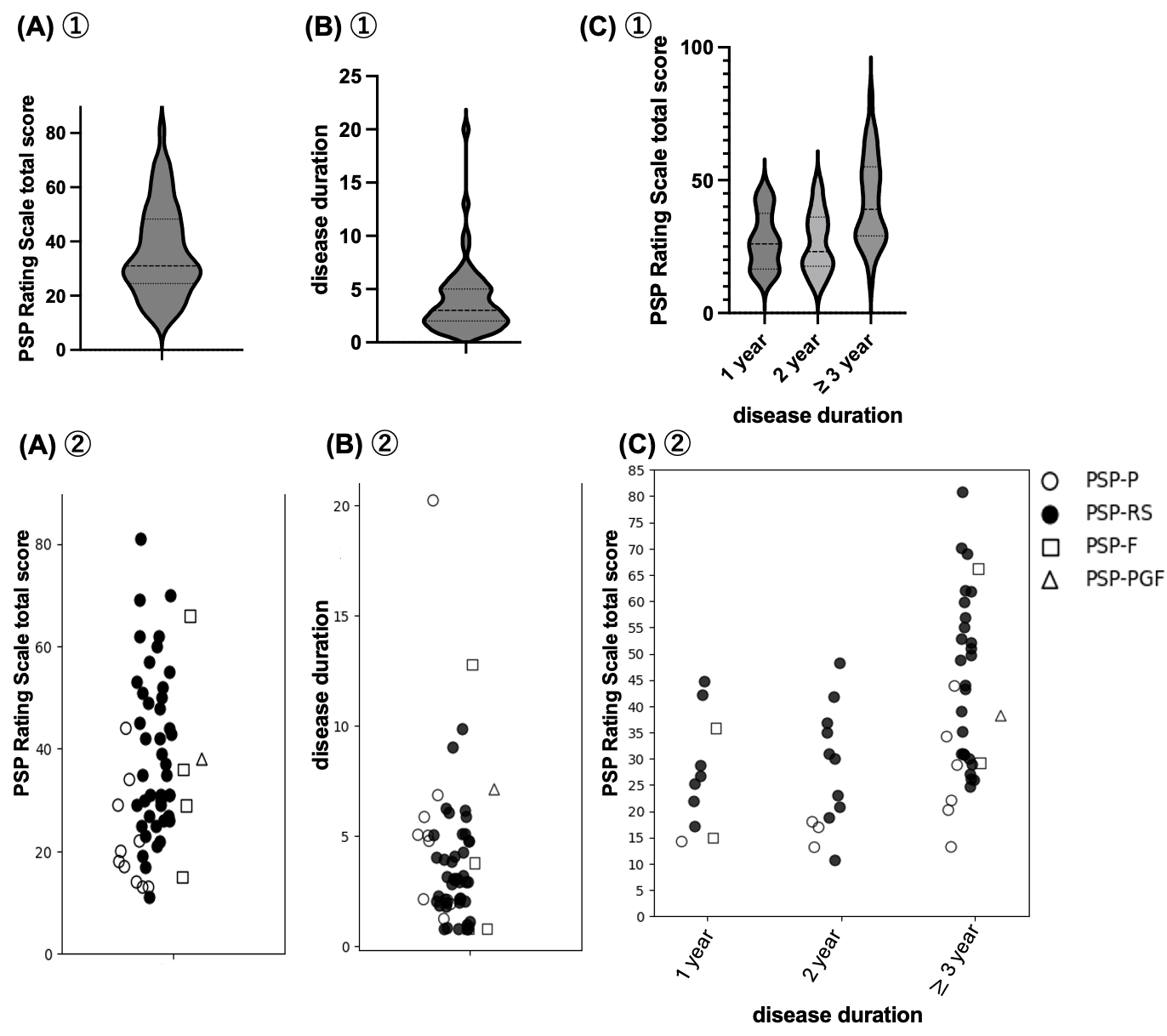

The distribution of disease duration and overall disease severity in the PSP patient cohort included in this study is presented.

In panels (A) ①, the distribution of overall disease severity based on total PSP Rating Scale scores is shown; in panels (B) ①, the distribution of disease duration among patients with PSP is illustrated; and in panels (C) ①, disease severity based on total PSP Rating Scale scores is depicted across three groups stratified by disease duration, using violin plots. Panels (A) ②, (B) ②, and (C) ② present the same data as in the corresponding ① panels, but with plots that allow differentiation of PSP subtypes.

This figure is intended to visualize the variability in disease stage and severity within the PSP cohort and to provide complementary information for interpreting the relationship between imaging findings and clinical features across different stages of disease. **eFigure 7: Distributions of tau accumulation (red), atrophy (green), and their overlaps (yellow) associated with neurological symptoms in PSP-RS.**

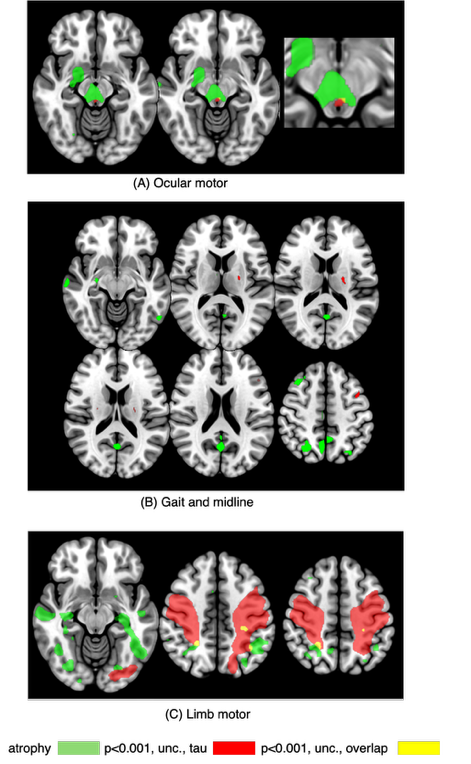

Analyses were restricted to patients with PSP-RS. Subtype classification was performed using the MAX rule, in which a diagnosis of PSP-RS was prioritized when applicable. Because the number of cases decreased to 43 when limited to PSP-RS, the following analyses were conducted as exploratory investigations.

First, in voxel-wise analyses restricted to PSP-RS, the distributions of tau accumulation and regional atrophy associated with neurological symptoms were broadly consistent with those observed in the overall PSP cohort; however, the spatial extent of these associations was more limited. Specifically, (A) ocular motor impairment was associated with tau accumulation and atrophy in the midbrain tegmentum, (B) gait and midline impairment with tau accumulation and atrophy primarily in the basal ganglia, and (C) limb motor impairment with tau accumulation mainly in the precentral gyrus and atrophy predominantly in the supramarginal gyrus (unc. *p <* 0.001).

RS, Richardson’s syndrome; unc., uncorrected.

**eFigure 8: Path analysis among tau probe retention, brain atrophy, and neurological symptoms in all patients with PSP-RS.**

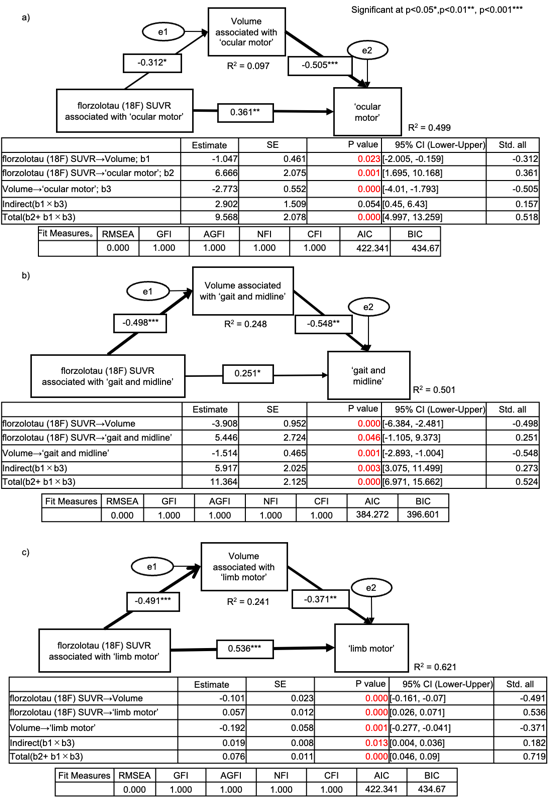

Path analysis was performed to examine hypothesized associations among tau tracer SUVR, regional brain volume, and symptom severity in the “ocular motor” (a), “gait and midline” (b), and “limb motor” symptom domains. Consistent with analyses in the overall PSP cohort, symptom severity was significantly associated with both regional brain volume and tau tracer SUVR, with stronger associations observed for regional brain volume.

RS, Richardson’s syndrome; SUVR, standardized uptake value ratio; SE, standard error; CI, confidence interval; Std. all, fully standardized coefficients; RMSEA, Root Mean Square Error of Approximation; GFI, Goodness‑of‑Fit Index; AGFI, Adjusted Goodness‑of‑Fit Index; NFI, Normed Fit Index; CFI, Comparative Fit Index; AIC, Akaike Information Criterion; BIC, Bayesian Information Criterion.

**eFigure 9: Path analysis among tau probe retention, brain atrophy, and neurological symptoms in PSP-RS patients classified by disease duration**

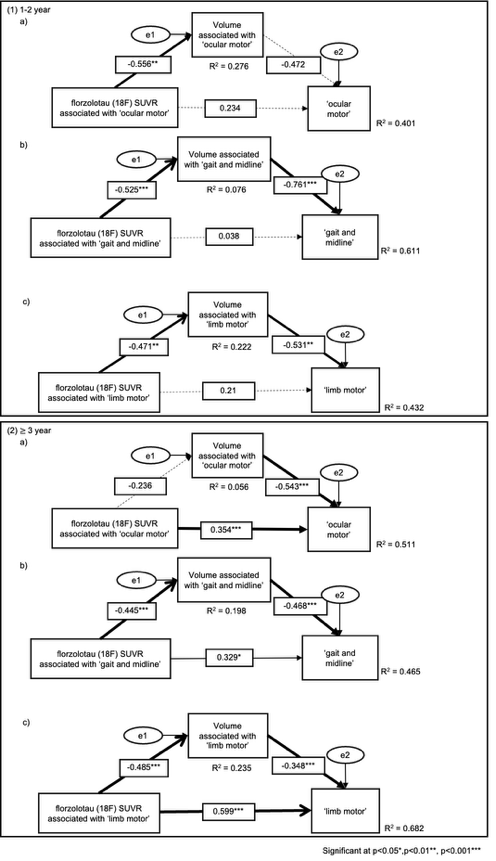

We stratified PSP-RS patients by disease duration. Because the subgroup with disease duration of 1 year or less was small (n = 7) and did not allow stable model estimation, we combined patients with disease duration of 1 to 2 years into an early-stage group (n = 17) and compared them with a late-stage group with disease duration of 3 years or longer (n = 26). Path analyses were then performed separately for each group for the ocular motor (a), gait and midline (b), and limb motor (c) symptom domains.

For the ocular motor domain, the early-stage group showed a relatively stronger association between tau burden and regional atrophy, whereas the late-stage group showed a more prominent association between regional atrophy and symptom severity. This pattern was similar to that observed in the analyses of the overall PSP cohort.

In contrast, for the gait and midline domain, the pattern differed from that in the overall PSP cohort. In the early-stage group, an association between regional atrophy and symptom severity was already evident in addition to the association between tau burden and regional atrophy. In the late-stage group, the association between regional atrophy and symptom severity was more pronounced.

RS, Richardson’s syndrome; SUVR, standardized uptake value ratio

**eFigure 10: Topology of tau accumulation across PSP subtypes**

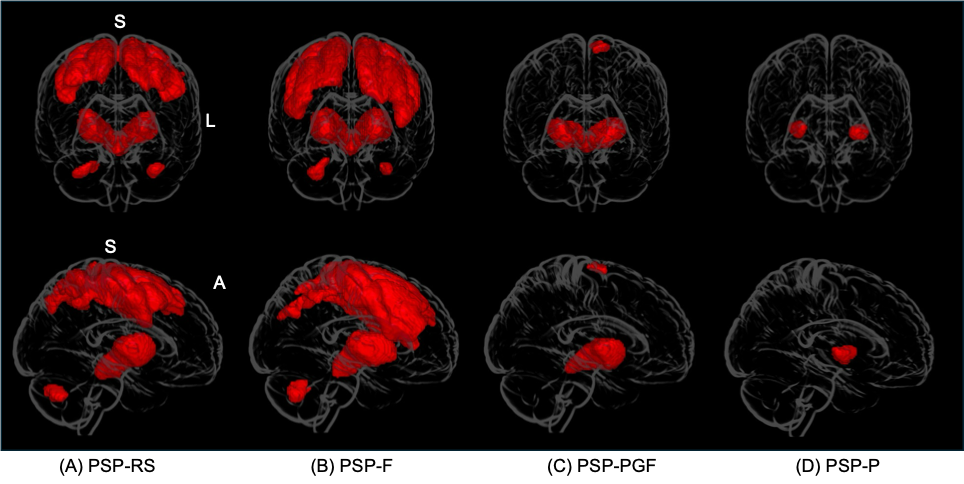

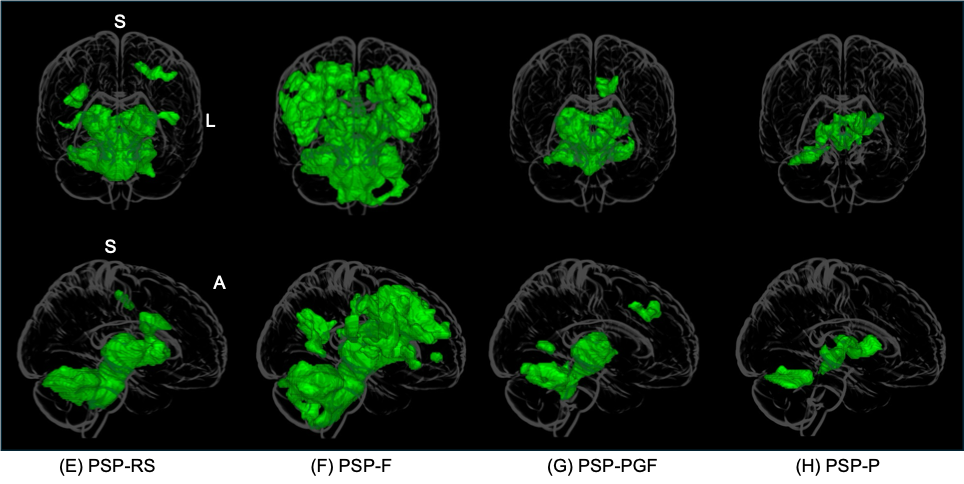

Voxel-wise analyses identified regions showing significantly increased tau accumulation in (A) PSP-RS, (B) PSP-F, (C) PSP-PGF, and (D) PSP-P compared with healthy controls. To evaluate subtype-specific distributions of tau accumulation, statistical maps were visualized at a voxel-level uncorrected threshold of *p <* 0.001, with only clusters exceeding the expected cluster size estimated by SPM being displayed. (*p <* 0.001, uncorrected; cluster size thresholds: k = 305 for (A), 255 for (B), 246 for (C), and 236 for (D)). In PSP-RS, tau accumulation was observed in the brainstem and basal ganglia and extended to the frontoparietal cortex. PSP-F showed a similar distribution, with more extensive involvement of the rostral middle frontal gyrus. In PSP-PGF, tau accumulation was predominantly localized to the midbrain and basal ganglia, whereas in PSP-P, it was largely confined to regions adjacent to the basal ganglia.

Voxel-wise analyses also identified regions with significantly reduced brain volume in (E) PSP-RS, (F) PSP-F, (G) PSP-PGF, and (H) PSP-P compared with healthy controls (*p <* 0.001, uncorrected; cluster size thresholds: k = 259 for (E), 185 for (F), 194 for (G), and 190 for (H)). In PSP-RS, atrophy was observed in the basal ganglia, cerebellum, and pons, whereas in PSP-F, atrophy extended to the frontal cortex in addition to these regions. In PSP-PGF, atrophy involved the basal ganglia and midbrain, while in PSP-P, it was localized to regions adjacent to the basal ganglia.

To capture a broader spectrum of clinical phenotypes, subtype classification was performed according to the MAX rule. When a subtype other than PSP-RS was considered the predominant clinical presentation, that subtype was preferentially assigned.

RS, Richardson’s syndrome; P, parkinsonism; F, frontal lobe cognitive or behavioral presentations; PGF, pure akinesia with gait freezing.

**eFigure 11: Tau deposition and atrophy regions associated with subdomain scores residualized for the total PSP Rating Scale score**

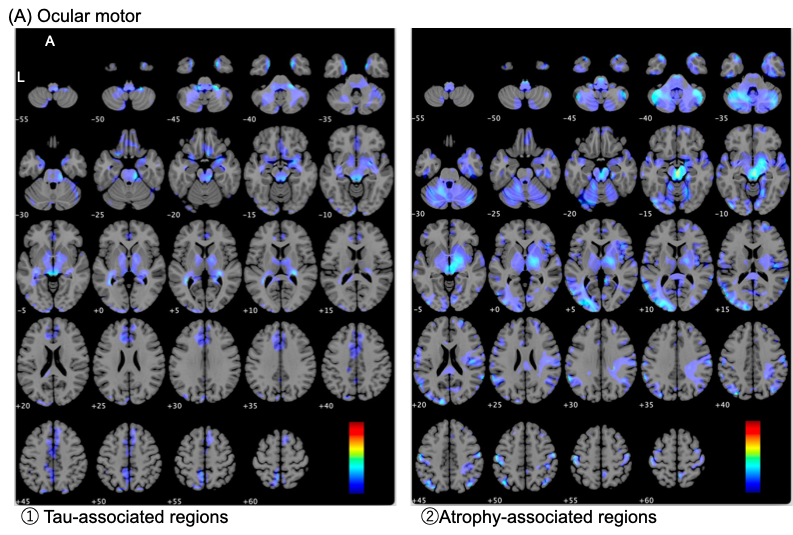

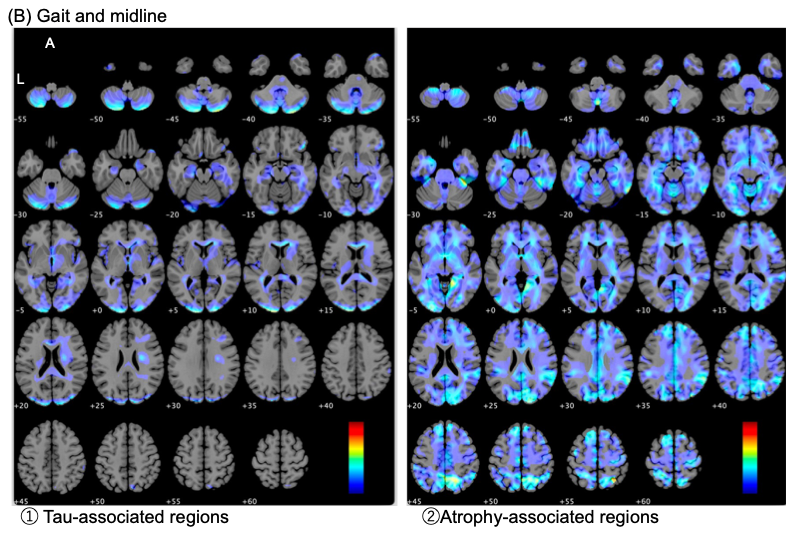

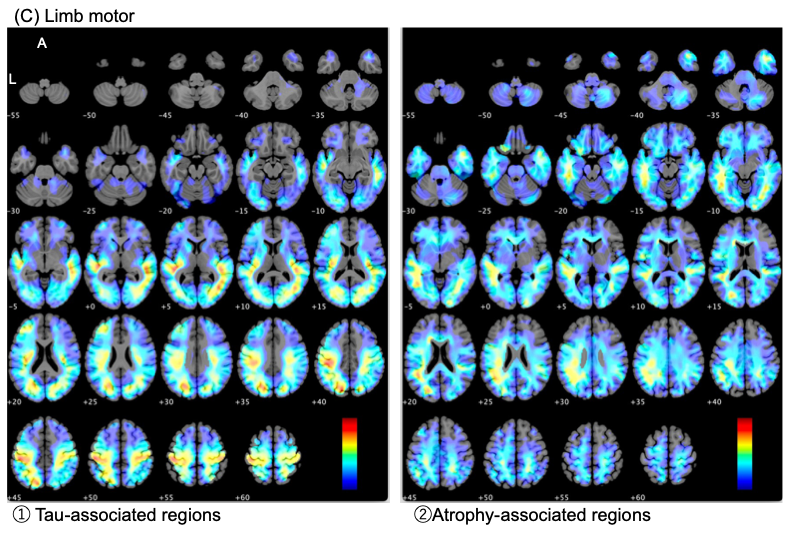

To examine domain-specific associations while minimizing the influence of overall disease severity, an exploratory analysis was conducted with the total PSP Rating Scale score included as a covariate. For visualization purposes, voxel-wise *t*-maps were displayed without applying a statistical threshold, using a color scale ranging from 0.5 to 5.5.

In this analysis, (A) regions of tau accumulation and atrophy associated with ocular motor symptoms were identified near the midbrain tegmentum. (B) In the gait and midline domain, few associations were observed in the basal ganglia. Instead, tau accumulation was associated with the cerebellar hemispheres, whereas atrophy was associated with the cerebellar vermis and occipital cortex. (C) In the limb motor domain, tau accumulation was observed in the precentral and postcentral gyri, as well as the occipital cortex, whereas atrophy extended from the supramarginal gyrus to the precentral and postcentral regions. Notably, regions associated with the limb motor domain retained clusters with relatively high t-values even after adjustment for the total PSP Rating Scale score.

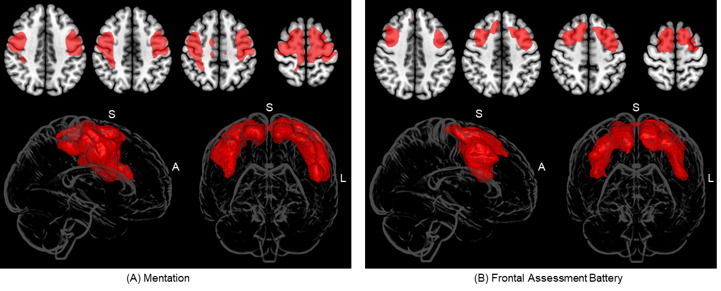
**eFigure 12: Distributions of tau accumulation (red) associated with neurological symptoms.**

(A) "Mentation" subscores of PSP Rating Scale scores associated with tau accumulation in the precentral gyrus and middle frontal gyrus (FWEc *p <* 0.05, k = 18486 voxels). (B) FAB scores associated with tau accumulation in the middle frontal gyrus and superior frontal gyrus (FWEc p < 0.05, k = 29346 voxels).

FAB, Frontal Assessment Battery.
