## Supplementary material for "*In Vivo* Topographic Associations Between Tau Pathology, Atrophy, and Symptom Domains in Patients with Progressive Supranuclear Palsy": eSAP

**Supplementary materials**

Supplementary material associated with this study can be found online.

Abbreviations: HC, healthy controls; PSP, progressive supranuclear palsy

**Study Protocol and Statistical Analysis Plan**

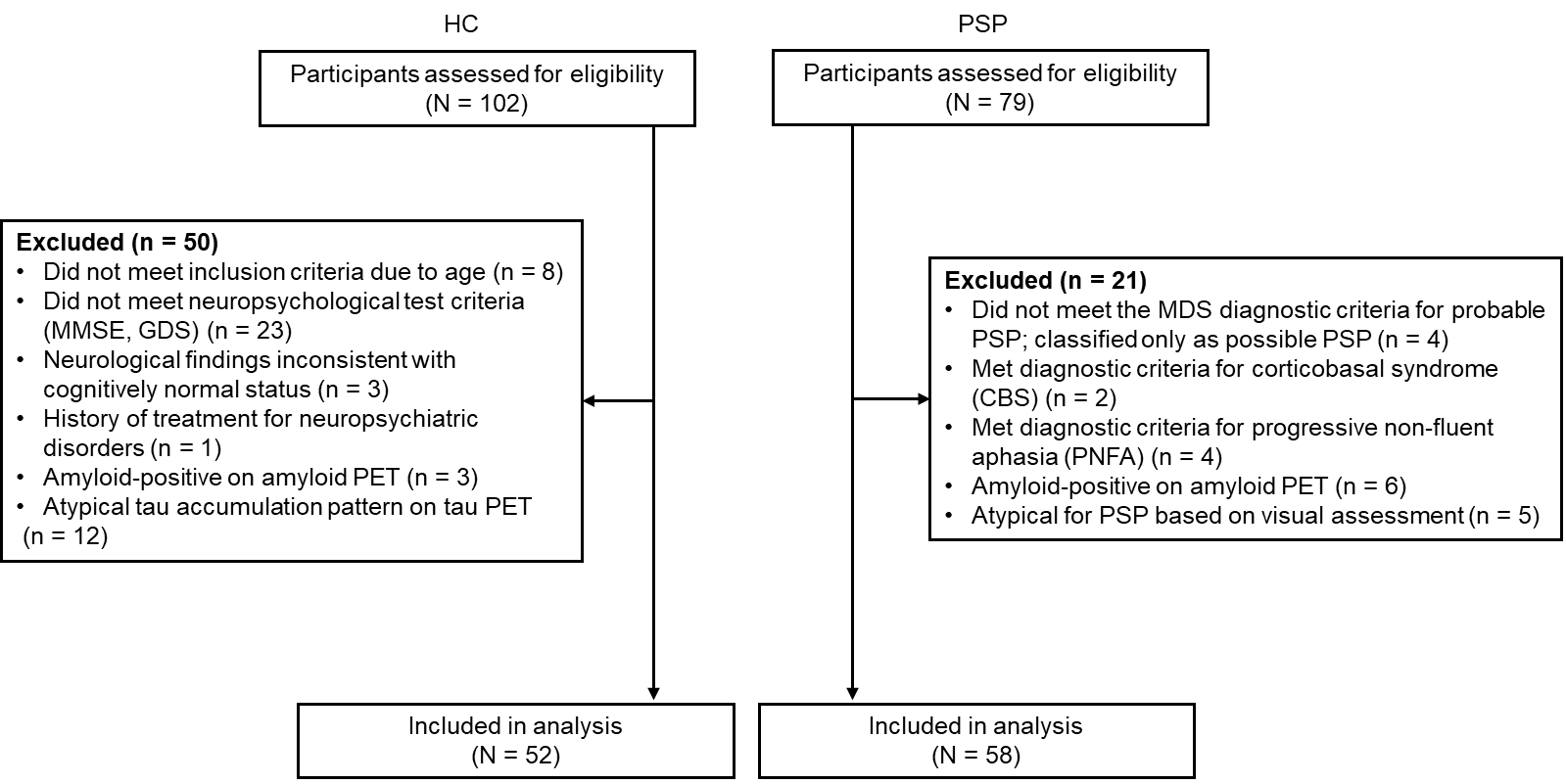
**eSAP 1: Trial flow diagram**

Among the 102 initially screened HC participants, 50 individuals were excluded for the following reasons. Twenty-three participants did not meet the neuropsychological inclusion criteria based on scores from the MMSE and the GDS. Eight individuals were excluded due to being outside the eligible age range. Twelve participants exhibited atypical tau accumulation patterns on tau PET imaging, and three were found to be amyloid-positive on amyloid PET. Additionally, three participants presented neurological findings inconsistent with a cognitively normal status, and one had a documented history of treatment for neuropsychiatric disorders.

In the progressive PSP group, 21 out of 79 screened patients were excluded. Specifically, four individuals did not meet the diagnostic criteria for PSP as defined by the Movement Disorder Society (MDS). Six patients were excluded due to amyloid positivity on amyloid PET. Two individuals met the clinical diagnostic criteria for CBS, and four were diagnosed with PNFA. Furthermore, five patients were considered atypical for PSP based on visual assessment of clinical or imaging findings. The remaining four patients were excluded due to insufficient data quality for reliable analysis.

MMSE, Mini-Mental State Examination; GDS, Geriatric Depression Scale; CBS; corticobasal syndrome; PNFA, progressive non-fluent aphasia.

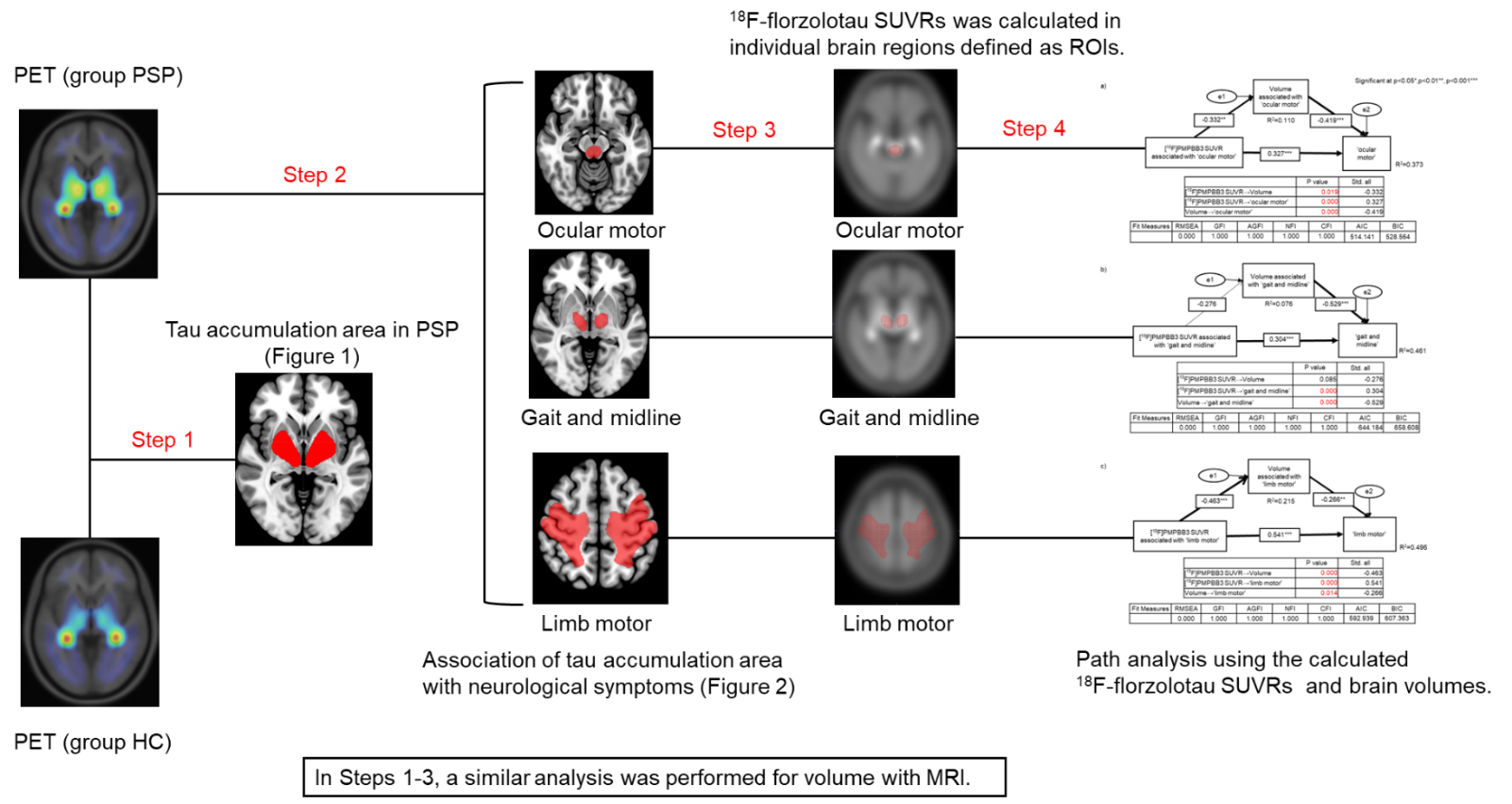
**eSAP 2: Research flow map**

A multistep analysis was performed to examine tau accumulation in PSP.

Step 1: Tau PET images were standardized, and voxel clusters showing significantly elevated tau probe retentions in PSP patients compared to HCs were identified.

Step 2: Voxel-wise analyses were conducted to determine voxel clusters where tau accumulation was significantly associated with neurological symptom severity.

Step 3: For each symptom domain, significant clusters identified in Step 2 were binarized to define ROIs, and SUVRs were calculated within these ROIs.

Similarly, Steps 1 - 3 were applied to structural MRI data to assess regional brain atrophy.

Step 4: Path analysis was performed using tau probe SUVRs and MRI-based regional brain volumes to model causal relationships.

ROI, regions of interest. SUVR, standardized uptake value ratio.
