## Supplementary material for "*In Vivo* Topographic Associations Between Tau Pathology, Atrophy, and Symptom Domains in Patients with Progressive Supranuclear Palsy": eMethods

**Supplementary materials**

Supplementary material associated with this study can be found online.

Abbreviations: HC, healthy controls; PSP, progressive supranuclear palsy

**eMethods**

*MRI Acquisition*

Structural MRI was performed using 3‑T MAGNETOM Verio or Prisma scanners (Siemens Healthcare). T1‑weighted gradient‑echo images were acquired using the following parameters: sagittal orientation; slice thickness, 1 mm; echo time [TE], 1.95 ms; repetition time [TR], 2,300 ms; flip angle, 9.0°; inversion time [TI], 900 ms; field of view [FOV], 250 mm; matrix size, 512 × 512 × 176. These images were used for PET coregistration and voxel‑based morphometry analyses.

*PET Acquisition and Image Reconstruction*

Amyloid pathology was evaluated using [11C]PiB PET. Scans were acquired on either a Biograph mCT Flow system (Siemens Healthcare; matrix dimension, 200 × 200 × 109; voxel size [mm], 2 × 2 × 2) or a Discovery MI scanner (GE Healthcare; matrix size: 128 × 128 × 89; voxel size: 2 × 2 × 2.8 mm³ for [11C]PiB scans). [11C]PiB images were acquired 50–70-mins after intravenous bolus injection (514.8 ± 82.6 MBq), typically in four 5-min frames.

Tau accumulation was assessed using florzolotau (18F) PET on a Biograph mCT Flow system (Siemens Healthcare; matrix dimension, 200 × 200 × 109; voxel size [mm], 2 × 2 × 2). List-mode event data were reconstructed using a filtered back-projection algorithm with a Hanning filter (6.0 mm full width at half maximum). Each participant underwent a 20-min PET acquisition (four 5-min frames or two 10-min frames) beginning 90 min after intravenous injection of florzolotau (18F) (193.4 ± 48.6 MBq). SUVR images were generated from motion-corrected florzolotau (18F) PET images acquired 90–110 min post-injection, using an automatically extracted gray matter (GM) reference region based on a signal histogram-based approach.

*Radioligand synthesis*

The radiosynthesis of florzolotau (18F) was performed as described elsewhere.^e1-3^ In brief, fluoride (18F) reacted with the tosylate precursor of florzolotau (18F) in the presence of dimethyl sulfoxide, K_2_CO_3_, and K222. The final formulated product of florzolotau (18F) was radiochemically pure (≥95%) as confirmed by analytical high-performance liquid chromatography (Waters Atlantis Prep T3 column, 4.6 × 150 mm; CH_3_CN/50 mM AcONH_4_ = 4/6, 1 ml/min).

*Data Preprocessing*

Data were preprocessed using PMOD 4.2 and 4.3 (PMOD Technologies LLC, Switzerland) and Statistical Parametric Mapping software (SPM12, Wellcome Department of Cognitive Neurology). Standardized uptake value ratio (SUVR) images were obtained from averaged PET images with motion correction at the following intervals: 90–110 min (florzolotau (18F)) post-injection, respectively. An in-house MATLAB (The MathWorks, Natick, MA, USA) script was implemented to automatically extract the gray matter (GM) reference region using a signal histogram-based approach for florzolotau (18F).^e4^ Each image (T1WI and florzolotau (18F) SUVR) was spatially normalized to the Montreal Neurological Institute Space (East Asian brain T1WI from International Consortium for Brain Mapping) using the Diffeomorphic Anatomical Registration Through Exponentiated Lie Algebra algorithm.

The SPM12 software was utilized for the exploratory voxel-based analysis of correlations among the PSP rating scale, each subscore with florzolotau (18F) SUVRs for tau pathology, and GM and white matter (WM) for volumes. All voxels outside the GM and WM masks were excluded from analysis (SPM Masking Toolbox^e5^) to avoid possible edge effects between different tissue types.

*Statistical and Path Analysis Procedures*

Independent-samples t-tests and χ2 tests were used to compare baseline demographic and clinical characteristics. All statistical tests were two-sided, with *p* < 0.05 considered statistically significant. Dat are presented as means ± standard deviations.

Demographic analyses were performed using EZR version 1.68 (Saitama Medical Center, Jichi Medical University, Saitama, Japan), a graphical user interface for R version 4.3.1 (The R Foundation for Statistical Computing, Vienna, Austria)^e6^, and Prism 9 for Windows 64-bit, version 9.5.1 (733) (GraphPad Software, Boston, USA).

Path analyses were performed using RStudio (version 2024.12.1+563, RStudio, PBC) and the R package lavaan^e7^ (version 0.6-16).

Path models were estimated using the robust maximum likelihood estimator (MLR). PSP Rating Scale subdomain scores were treated as ordinal variables because of their inherently ranked distributions. MLR was used to obtain parameter estimates robust to non-normality. Tau deposition was specified as the independent variable, atrophy as the mediator, and symptom-domain score as the dependent variable. Direct and indirect effects were evaluated.

Model fit was assessed using the normed fit index, comparative fit index, and root mean square error of approximation in all patients with PSP. Akaike’s information criterion was used to identify the preferred model.
