## Supplementary material for "*In Vivo* Topographic Associations Between Tau Pathology, Atrophy, and Symptom Domains in Patients with Progressive Supranuclear Palsy": eAppendix

**Supplementary Materials**

**eAppendix**

**eAppendix 1. Supplemental Discussion**

**Interpretation of tau accumulation and atrophy distributions in PSP-RS**

As shown in eFigure 7, when the analysis was restricted to PSP‑RS, the regions exhibiting significant associations were highly limited. As illustrated in eFigure 3, this may be attributable to the restriction to PSP‑RS cases, which likely resulted in a cohort biased toward more advanced disease, thereby reducing the detectability of such associations.

RS, Richardson’s syndrome.

**eAppendix 2. Supplemental Discussion**

**Interpretation of path analyses in PSP-RS stratified by disease duration**

As shown in eFigure 9, we performed path analyses in PSP‑RS cases further stratified into early and late disease-duration groups to examine relationships among tau deposition, atrophy, and neurological symptoms. In the "gait and midline" domain, atrophy exhibited a relatively prominent association with clinical symptoms even in the early-stage group.

These findings may reflect the inclusion of cases within the first and second years after disease onset in the early-stage group, as well as the restriction to PSP‑RS cases, which may have led to earlier manifestation of the impact of progressive atrophy.

RS, Richardson’s syndrome.

**eAppendix 3. Supplemental Discussion**

**Interpretation of the topology of tau accumulation and atrophy across PSP subtypes**

In eFigure 10, to visualize and evaluate more diverse patterns of tau accumulation and atrophy across PSP subtypes, classification was performed based on the MAX rule ^e8^, whereby cases were assigned to alternative dominant subtypes when non PSP-RS features were considered predominant. Although the MDS‑PSP subtype classification is based on clinical features at the time of assessment, subtypes may evolve over the course of disease progression, and the presence of multiple clinical features may introduce interpretive variability in classification. Nevertheless, the distribution patterns of tau accumulation and brain atrophy observed in this analysis were consistent with the clinical characteristics of each subtype.

**eAppendix 4. Supplemental Discussion**

**Interpretation of tau deposition and atrophy regions associated with subdomain scores residualized for the total PSP Rating Scale score**

After residualizing for the total PSP Rating Scale score, we examined regional associations of tau accumulation and atrophy with neurological symptoms. As shown in eFigure 11, in the ocular motor domain, a subset of regions identified before residualization remained, although the extent of these associations was reduced. In contrast, in the gait and midline domain, most of the previously associated basal ganglia regions regions were no longer significant after residualization. These findings suggest that associations in the ocular motor domain are not entirely explained by overall disease severity, whereas those in the gait and midline domain appear to be more strongly driven by global disease severity.

In the limb motor domain, regions of tau accumulation and atrophy remained relatively widespread even after adjustment for total scores. This finding suggests that cortical tau deposition and atrophy more strongly reflect interindividual variability independent of overall disease severity.

**eAppendix 5. Supplemental Discussion**

**Supplementary Discussion on Remote Neurodegeneration**

Although prior neuropathological studies have reported interneuron loss in the primary motor cortex alongside extensive glial and neuronal tau inclusions, ^e9^ the relative preservation of large pyramidal neurons may mitigate detectable atrophy in this region. This notion aligns with a previous report documenting sparing of the motor cortex volume in PSP-Richardson type. ^e10^ Conversely, degeneration of cortico-cortical projection neurons was identified in the pre-supplementary motor area, ^e9^ and tau-induced axonal pathology may also disrupt intercortical pathways originating from the primary motor cortex. These degenerative processes could impair the delivery of trophic support from axonal terminals to downstream regions, such as the angular gyrus, ultimately leading to transneuronal degeneration and atrophy in areas remote from dense tau accumulations.

**eAppendix 6. Supplemental Discussion**

**Supplementary Discussion on Neocortical Contributions to Gait and Midline Symptoms**

In addition, tau deposition and atrophy in distinct neocortical regions were also associated with "gait and midline" symptoms, indicating that tau-associated neuronal loss in remote areas may contribute to these deficits. Tau accumulation in the lateral occipital cortex, including the secondary visual cortex (V_2_), may disrupt visual processing relevant to locomotion, ^e11^ while atrophy in the precuneus may compromise sensory integration required for spatiotemporal control of motor behavior. ^e11,e12^ Given the known functional connectivity between V_2_ and the dorsal precuneus, ^e13^ tau deposition in V_2_ may adversely affect neuronal viability in the precuneus via axonal projections. Alternatively, tau pathology in the thalamus may impact the precuneus through its strong structural and functional connectivity. ^e14^

**eAppendix 7. Supplemental Discussion**

**Supplementary Discussion on Neocortical Contributions to Limb Motor Dysfunction**

Prior studies have demonstrated activation of the fronto-temporo-parietal network during pantomime and imitation tasks, mediated by the superior longitudinal fasciculus, supporting its role in apraxia and its association with temporal and parietal degeneration. ^e15^

Our findings further suggest that tau accumulation in the primary motor and primary somatosensory cortices may exert secondary effects on neuronal integrity in downstream projection sites. In this context, the superior longitudinal fasciculus, which connects the primary motor cortex and angular gyrus, ^e16^ may serve as a conduit for transneuronal degeneration, ultimately contributing to volume loss in parietal regions.

Moreover, the motor cortex and parietal lobe, including the angular gyrus, are considered to form a sensory–motor integration network, and dysfunction within this network may contribute to dystonia, a key feature of the limb motor domain. ^e17^

**eAppendix 8. Supplemental Discussion**

**Interpretation of negative findings in the history, mentation, and bulbar domains**

In the voxel-wise association analyses, we did not identify significant clusters linking tau PET retention or regional brain volume differences to PSP Rating Scale subscores in the history, mentation, or bulbar domains. Given the clinical relevance of bulbar dysfunction in PSP, these negative findings warrant explicit consideration.

**History domain:**

The history domain encompasses multidimensional clinical features, including neuropsychiatric symptoms, dysphagia, motor impairments of the limbs and trunk, sleep disturbances, and urinary dysfunction. The diversity of these components and the difficulty in attributing them to specific anatomical substrates may have influenced the observed associations.

**Mentation domain:**

The mentation domain includes not only frontal lobe functions but also orientation. While it showed associations with frontal tau accumulation, no corresponding atrophy was detected, suggesting a potential dissociation between tau deposition and structural changes in this domain.

**Bulbar domain:**

The bulbar domain of the PSP Rating Scale comprises only two items, inherently limiting score granularity and dynamic range. In addition, quantitative assessment of the lower brainstem using PET and MRI is technically challenging, which may have constrained the sensitivity of MRI and static molecular PET measures to fully capture relevant pathological changes.
