## Supplementary material for "*In Vivo* Topographic Associations Between Tau Pathology, Atrophy, and Symptom Domains in Patients with Progressive Supranuclear Palsy": eTable

**eTable1: Clinical characteristics of PSP subtypes included in the analysis shown in eFigure 7**

|  | | **PSP subtype** | | | | **P-value** |
| --- | --- | --- | --- | --- | --- | --- |
|  |  | **PSP-RS　(n = 43)** | **PSP-F　(n = 4)** | **PSP-PGF　(n = 1)** | **PSP-P　(n = 10)** |  |
| Women, n (%) | | 18 (41.9) | 1 (25.0) | 1 (100.0) | 5 (50.0) | 0.557 |
| Age (years) | | 71.47 (7.94) | 73.00 (2.94) | 80.00 (NA) | 70.80 (8.15) | 0.599 |
| Disease duration (years) | | 3.37 ± 2.06 | 4.75 ± 5.68 | 7.0 | 5.5 ± 5.48 | 0.307 |
| PSP Rating Scale total(point) | | 39.35 ± 16.05** | 36.5 ± 21.52 | 38.0 | 22.4 ± 10.25 | 0.016 |
|  | History (point) | 9.5 ± 4.7* | 8.3 ± 6.9 | 9.0 | 5.0 ± 2.5 | 0.033 |
|  | Mentation (point) | 3.1 ± 3.4 | 3.0 ± 1.8 | 5.0 | 0.9 ± 1.1 | 0.066 |
|  | Bulbar (point) | 3.0 ± 2.0 | 4.3 ± 1.3 | 1.0 | 1.7 ± 1.3 | 0.069 |
|  | Ocular motor (point) | 7.7 ± 3.7* | 8.3 ± 5.4 | 7.0 | 4.4 ± 2.8 | 0.045 |
|  | Limb motor (point) | 4.4 ± 2.6 | 4.5 ± 1.0 | 4.0 | 4.0 ± 1.6 | 0.943 |
|  | Gait and midline (point) | 11.6 ± 4.6* | 8.3 ± 6.7 | 12.0 | 6.4 ± 4.0 | 0.021 |
| FAB (point) | | 11.4 ± 4.2* | 8.5 ± 1.3** | 14.0 | 15.4 ± 1.8 | 0.004 |
| Data are expressed as the number (%) or mean ± SD, as appropriate; age, disease duration, PSP Rating Scale total and subdomain scores, and FAB score are expressed as mean ± SD. | | | | | |  |
| ^**^P < 0.01, ^*^P < 0.05 versus PSP-P group post hoc Dunn’s test | | | | |  |  |

Clinical characteristics of patients with PSP-RS, PSP-F, PSP-PGF, and PSP-P included in the analysis presented in eFigure 7.

FAB, Frontal Assessment Battery; PSP-F, PSP with predominant frontal presentation; PSP-PGF, PSP with progressive gait freezing; PSP-P, PSP-parkinsonism; PSP-RS, PSP-Richardson syndrome.

**eTable 2: Clinical characteristics of PSP subtypes included in the analysis shown in eFigure 10**

|  | | **PSP subtype** | | | | | **P-value** |
| --- | --- | --- | --- | --- | --- | --- | --- |
|  |  | **PSP-RS　(n = 25)** | **PSP-F　(n = 13)** | **PSP-PGF　(n = 10)** | **PSP-P　(n = 10)** | |  |
| Women, n (%) | | 11 (44.0) | 5 (38.5) | 4 (40.0) | 5 (50.0) | | 0.949 |
| Age (years) | | 72.8 ± 7.3 | 71.5 ± 7.3 | 69.6 ± 9.3 | 70.8 ± 8.2 | | 0.725 |
| Disease duration (years) | | 3.1 ± 2.2 | 3.5 ± 3.1 | 4.7 ± 2.3 | 5.5 ± 5.5 | | 0.142 |
| PSP Rating Scale total(point) | | 36.7 ± 18.2^*^/100 | 43.6 ± 16.2^**^/100 | 39.1 ± 9.1^*^/100 | 22.4 ± 10.3/100 | | 0.005 |
|  | History (point) | 8.8 ± 4.7/24 | 10.6 ± 5.6^*^/24 | 9.3 ± 3.7/24 | 5.0 ± 2.5/24 | | 0.021 |
|  | Mentation (point) | 2.6 ± 3.3^#^/16 | 5.2 ± 3.3^**^/16 | 1.6 ± 1.7^#^/16 | 0.9 ± 1.1/16 | | 0.001 |
|  | Bulbar (point) | 2.6 ± 1.9/8 | 3.9 ± 1.9^*^/8 | 3.1 ± 2.0/8 | 1.7 ± 1.3/8 | | 0.061 |
|  | Ocular motor (point) | 7.6 ± 3.8/16 | 8.2 ± 3.9/16 | 7.7 ± 3.6/16 | 4.4 ± 2.8/16 | | 0.043 |
|  | Limb motor (point) | 4.6 ± 3.2/16 | 4.1 ± 1.3/16 | 4.3 ± 1.6/16 | 4.0 ± 1.6/16 | | 0.931 |
|  | Gait and midline (point) | 10.4 ± 5.2/20 | 11.8 ± 4.9/20 | 13.1 ± 2.4^*^/20 | 6.4 ± 4.0/20 | | 0.012 |
| FAB (point) | | 12.4 ± 3.9^#^/18 | 7.9 ± 4.0^****^/18 | 12.5 ± 2.3/18 | 15.4 ± 1.8/18 | | <0.001 |
| Data are expressed as the number (%) or mean ± SD, as appropriate; age, disease duration, PSP Rating Scale total and subdomain scores, and FAB score are expressed as mean ± SD. | | | | | | |  |
| ^****^P < 0.0001, ^**^P < 0.01, ^*^P < 0.05 versus PSP-P group post hoc Dunn’s test | | | | | |  |  |
| ^##^P < 0.01, ^#^P < 0.05 versus PSP-F group post hoc Dunn’s test | | | | | |  |  |

Clinical characteristics of patients with PSP-RS, PSP-F, PSP-PGF, and PSP-P included in the analysis presented in eFigure 11.

The total PSP Rating Scale score was significantly higher in PSP-RS, PSP-F, and PSP-PGF than in PSP-P. Compared with PSP-P, PSP-F showed significantly higher mentation and history subscores and a significantly lower FAB score, whereas PSP-PGF exhibited a significantly higher gait and midline subscore. Although the differences did not reach statistical significance, PSP-RS tended to show relatively higher ocular motor and gait and midline subscores.

FAB, Frontal Assessment Battery; PSP-F, PSP with predominant frontal presentation; PSP-PGF, PSP with progressive gait freezing; PSP-P, PSP-parkinsonism; PSP-RS, PSP-Richardson syndrome.
