## Supplementary material for "*In Vivo* Topographic Associations Between Tau Pathology, Atrophy, and Symptom Domains in Patients with Progressive Supranuclear Palsy": eReference

eMethods

e1.	Kimura Y, Ichise M, Ito H, et al. PET Quantification of Tau Pathology in Human Brain with 11C-PBB3. *J Nucl Med*. Sep 2015;56(9):1359-65. doi:10.2967/jnumed.115.160127

e2.	Maruyama M, Shimada H, Suhara T, et al. Imaging of tau pathology in a tauopathy mouse model and in Alzheimer patients compared to normal controls. *Neuron*. Sep 18 2013;79(6):1094-108. doi:10.1016/j.neuron.2013.07.037

e3.	Ridgway GR, Omar R, Ourselin S, Hill DL, Warren JD, Fox NC. Issues with threshold masking in voxel-based morphometry of atrophied brains. *Neuroimage*. Jan 1 2009;44(1):99-111. doi:10.1016/j.neuroimage.2008.08.045

e4.	Tagai K, Ikoma Y, Endo H, et al. An optimized reference tissue method for quantification of tau protein depositions in diverse neurodegenerative disorders by PET with (18)F-PM-PBB3 ((18)F-APN-1607). *Neuroimage*. Dec 1 2022;264:119763. doi:10.1016/j.neuroimage.2022.119763

e5.	Tagai K, Ono M, Kubota M, et al. High-Contrast In Vivo Imaging of Tau Pathologies in Alzheimer's and Non-Alzheimer's Disease Tauopathies. *Neuron*. Jan 6 2021;109(1):42-58 e8. doi:10.1016/j.neuron.2020.09.042

e6.	Gan L, Yan R, Su D, et al. Alterations of structure and functional connectivity of visual brain network in patients with freezing of gait in Parkinson's disease. *Front Aging Neurosci*. 2022;14:978976. doi:10.3389/fnagi.2022.978976

e7.	Rosseel Y. lavaan: An R Package for Structural Equation Modeling. *J Stat Softw*. 05/24 2012;48(2):1 - 36. doi:10.18637/jss.v048.i02

e8.	Grimm MJ, Respondek G, Stamelou M, et al. How to apply the movement disorder society criteria for diagnosis of progressive supranuclear palsy. *Mov Disord*. Aug 2019;34(8):1228-1232. doi:10.1002/mds.27666

e9.	Halliday GM, Macdonald V, Henderson JM. A comparison of degeneration in motor thalamus and cortex between progressive supranuclear palsy and Parkinson's disease. *Brain*. Oct 2005;128(Pt 10):2272-80. doi:10.1093/brain/awh596

e10.	Whitwell JL, Tosakulwong N, Botha H, et al. Brain volume and flortaucipir analysis of progressive supranuclear palsy clinical variants. *Neuroimage Clin*. 2020;25:102152. doi:10.1016/j.nicl.2019.102152

e11.	Bommarito G, Putzolu M, Avanzino L, et al. Functional Correlates of Action Observation of Gait in Patients with Parkinson's Disease. *Neural Plast*. 2020;2020:8869201. doi:10.1155/2020/8869201

e12.	Cavanna AE, Trimble MR. The precuneus: a review of its functional anatomy and behavioural correlates. *Brain*. Mar 2006;129(Pt 3):564-83. doi:10.1093/brain/awl004

e13.	Zhang S, Li CS. Functional connectivity mapping of the human precuneus by resting state fMRI. *Neuroimage*. Feb 15 2012;59(4):3548-62. doi:10.1016/j.neuroimage.2011.11.023

e14.	Cunningham SI, Tomasi D, Volkow ND. Structural and functional connectivity of the precuneus and thalamus to the default mode network. *Hum Brain Mapp*. Feb 2017;38(2):938-956. doi:10.1002/hbm.23429

e15.	Vry MS, Tritschler LC, Hamzei F, et al. The ventral fiber pathway for pantomime of object use. *Neuroimage*. Feb 1 2015;106:252-63. doi:10.1016/j.neuroimage.2014.11.002

e16.	Wang X, Pathak S, Stefaneanu L, Yeh FC, Li S, Fernandez-Miranda JC. Subcomponents and connectivity of the superior longitudinal fasciculus in the human brain. *Brain Struct Funct*. May 2016;221(4):2075-92. doi:10.1007/s00429-015-1028-5

e17.	Perruchoud D, Murray MM, Lefebvre J, Ionta S. Focal dystonia and the Sensory-Motor Integrative Loop for Enacting (SMILE). *Front Hum Neurosci*. 2014;8:458. doi:10.3389/fnhum.2014.00458
